# Pharmacogenetic phenoconversion modeling of drug–drug–gene interactions on CYP2C19 activity: effects of comedication by genotype on escitalopram concentrations

**DOI:** 10.64898/2026.06.23.26356327

**Authors:** Julia Stingl, Espen Molden, Kristine Hole, Birgit M. Wollmann, Roberto Viviani

## Abstract

**Background**. Polypharmacy is an important source of phenoconversion caused by drug interactions potentially modulated by genetic variability. **Aims**. To develop a linear phenoconversion model for TDM data and provide quantitative estimates of drug-drug-gene interactions (DDGIs) in the pharmacogenetic phenotype groups of CYP2C19. **Methods**. Escitalopram TDM data in a large real-world sample (n=2,852) was analysed for phenoconversion of CYP2C19 activity. Co-medication was identified by reprocessing high-resolution mass-spectra (Orbitrap). We developed a statistical model to identify inhibition from co-medication in the CYP2C19 and in alternative elimination pathways. We extended the model to estimate the inhibition ensuing from individual co-medications, using a single model for all data to account for multiple co-medications and confounders simultaneously. A Bayesian approach allowed us to stabilize the fit and provide well-calibrated credibility intervals. **Results**. Reprocessing of TDM analyses identified 17 co-medications, which were shown to phenoconvert CYP2C19 activity proportionally to the activity in non-medicated phenotypes. Phenoconversion decreased the original CYP2C19 activity by about one third for a co-medication that corresponded to a 100% substrate of CYP2C19. The extent of CYP2C19 phenoconversion correlated strongly with the fractional contribution of CYP2C19 to the metabolism of the specific co-medication reported in the pharmacogenetic literature (R^2^=0.55) so long as the mechanism was competitive inhibition. **Conclusion**. We provide the statistical methodology to estimate phenoconversion from co-medication in TDM data and combine TDM and pharmacogenetic datasets in future studies aiming at establishing quantitative models of DDGIs.

## Introduction

Despite the availability of robust pharmacogenetic dosing guidelines for many psychotropic drugs, their clinical utility remains constrained by a critical gap: current approaches insufficiently account for the dynamic impact of co-medication on drug-metabolizing enzymes, particularly under conditions of polypharmacy that are common in psychiatric practice. As a result, genotype-based predictions frequently fall short of capturing real-world variability in drug exposure (1). This challenge reflects a broader limitation in psychopharmacology, where pharmacogenetic and drug–drug interaction effects are still largely considered separately rather than within a unified framework (2).

Clinical decision-support systems and drug interaction databases typically generate warnings for drug combinations that compete for metabolism via the same enzymatic pathway (3). However, these alerts rarely capture the complex reality of polypharmacy, where multiple drugs may simultaneously compete for or inhibit the same enzyme. In real-world clinical practice, patients’ medication regimens may present virtually unique co-medication combinations (4). As a result, potential drug-drug interactions can differ substantially between patients and may change over time as therapies are adjusted. Importantly, most concomitant medications encountered in routine practice act as competitive or weak inhibitors rather than as strong inhibitors, underscoring the need for a nuanced and context-dependent assessment rather than a binary classification of interaction risk.

The ability to quantifiably predict specific drug-drug interactions may further be complicated by individual susceptibility towards metabolic changes related to genetic variability of drug-metabolizing enzymes caused by genetic polymorphisms, as well as from drug interactions that alter enzyme function — a process known as phenoconversion (5). The combined influence of genetic variability and pharmacokinetic drug-drug interactions has been conceptualized as drug-drug-gene interactions (DDGIs) (6). Despite growing awareness of their clinical importance, few studies have provided quantitative estimates of phenoconversion under real-world conditions (7).

In the present study, we used statistical methodology to quantify the degree of phenoconversion of CYP2C19 activity in a large real-world population of *CYP2C19*-genotyped patients medicated with escitalopram from the Norwegian therapeutic drug monitoring (TDM) database on psychiatric drugs (8). Escitalopram is a major CYP2C19 substrate, with minor contributions of CYP2D6 and CYP3A4 (9). Reprocessing of high-resolution mass spectra by exact mass and fragment detection (Orbitrap) identified possible co-medication (10). This approach yielded individual co-medication profiles that were then used to estimate the inhibitory effects on CYP2C19 activity, integrated with CYP2C19 pharmacogenetic data from 2,852 individuals.

Although several models have been proposed in the literature, empirical evidence suggests that a simple linear phenoconversion model may adequately approximate inhibition effects despite its conceptual simplifications (7). Here, our first aim was to adapt this model to TDM data and apply it to a large polypharmacy dataset to estimate phenoconversion quantitatively (6) (Figure 1).

**Figure 1.**
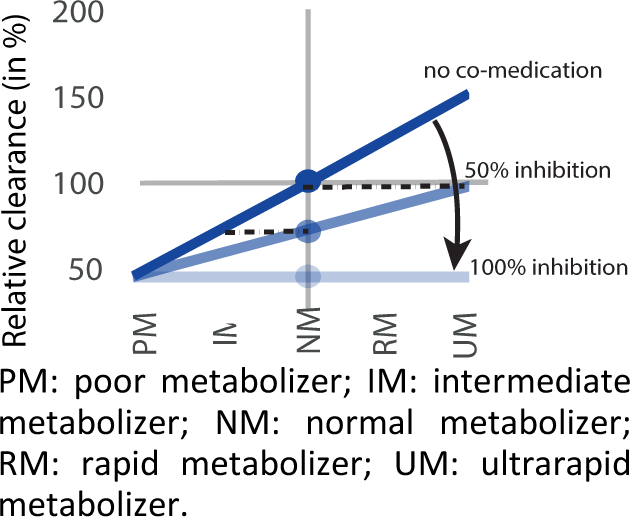
In the linear phenoconversion model, a straight line representing increasing clearance ratios for higher metabolism phenotype groups (blue line). This line progressively flattens to the level of poor metabolizer activity with increasing extent of enzyme inhibition. Under the model, the relationship remains linear because inhibition is proportional to the original clearance levels. The horizontal dashed lines exemplify the phenoconversion. In the hypothetical example of the Figure, 50% inhibition converts ultrarapid metabolizers to normal metabolizers, and normal metabolizers to intermediate metabolizer phenotypes (adapted from (6)).

The pharmacogenetic literature provides estimates of the fractional contribution of the polymorphic enzyme to the metabolism of individual drugs based on the comparison of clearance levels in normal and poor metabolizers. While these estimates provide a rationale for dose adjustments in carriers of the variant phenotypes of the drugs in question, it is less clear how well they may predict the effect of this drug on the metabolism of a second drug, i.e. the magnitude of interactions due to competitive inhibition of an enzyme metabolizing multiple substrates administered simultaneously (2). Hence, the second aim of the study was to assess the capacity of the phenoconversion model to predict the interaction magnitude in real-world data from pharmacogenetic fractional contribution estimates. We will show this to be the case for interactions arising from competitive inhibition, while other interaction mechanisms provided datapoints that clearly deviated from the prediction.

## Methods

### Escitalopram serum concentration and genotyping

A dataset with serum samples of n=2,873 escitalopram-medicated patients was retrieved from the Norwegian TDM database at the Center for Psychopharmacology, Diakonhjemmet Hospital in Oslo. Concentrations of escitalopram from serum samples as well as the major metabolite N-desmethyl escitalopram were determined as a part of routine TDM (during 2021-2025) using a Ultra-High Performance Liquid Chromatography-High-Resolution (Orbitrap) Mass Spectrometry system, as previously described (10). Briefly, chromatographic separation was achieved using an XBridge BEH C18 column (2.5 µm, 2.1 · 75 mm) (Waters Corporation, Milford, MA, US), where a run time of 3.5 minutes enabled detection of both compounds.

In parallel, pharmacogenetic analysis was conducted in *CYP2C19* to determine the alleles *CYP2C19*2, *3, *4, *17*. These allelic diplotypes were used to predict poor metabolizer (PM), intermediate metabolizer (IM), normal metabolizer (NM) and ultrarapid metabolizer (UM) phenotypes.

### Data preparation and sample description

A series of preliminary analyses were conducted to screen for potential confounders and the existence of outliers (see Supplementary file 1). These models identified one extreme outlier and 20 individuals with very high escitalopram clearance but also low plasma concentrations of the metabolite, suggesting that they were not taking the medication at the prescribed doses. These individuals were excluded from the analysis, giving a final sample size of n=2,852 (mean age 45.75, range 14-101, 64.3% females, escitalopram prescribed dose mode 10 mg, range 2.5-50 mg). More details on the sample are in Supplementary file 1.

### Relationship between CYP2C19 clearance and escitalopram serum concentration data

As a proxy of escitalopram clearance values, we used the reciprocal of escitalopram concentration to dose ratios at trough level (D/C ratios at trough). These ratios have the dimensionality of volume over dosing interval (here, kiloliters per day, kL/day). Differences in D/C ratios may be used as a proxy of clearance differences based on the following assumptions:

- Under steady-state conditions and given the long half-life of escitalopram trough concentrations were assumed to approximate average steady-state exposure data.
- Linear pharmacokinetics were assumed, implying that plasma concentrations are proportional to dose, while clearance remains dose-independent (11).

It should be noted that D/C ratios derived from TDM data may be influenced by factors such as sampling variability, adherence, and prior dose adjustments. Non-adherence was addressed by inspecting the concentration of escitalopram and its metabolite and excluding participants whose concentration was low in both, a pattern suggesting lack of ingestion (see Supplementary file 1). The other factors were considered as confounders in the model.

### 11Identification of co-medication by Orbitrap high-resolution mass spectrometry

To obtain data on co-medication that could interact with CYP2C19-mediated escitalopram metabolism, retrospective reprocessing of Orbitrap high-resolution mass spectra from analyses of the respective escitalopram TDM samples was performed, thereby addressing the problem of the lack of information regarding co-prescribed drugs on the TDM request forms. We first produced a list of potentially interacting CYP2C19 substrates with available pharmacogenetic evidence (12) and restricted it to agents considered clinically relevant in Norway (n = 23). The list also included a few compounds in whose metabolism CYP2C19 plays little or no known role but were well represented in the reference library. These compounds represented implicit baseline controls in the regression, thereby improving statistical power. Based on this curated set, a systematic reprocessing of existing mass spectrometry data from patient blood samples was conducted to identify the presence of these compounds. The investigated substances comprised amitriptyline, clomipramine, doxepin, imipramine, lamotrigine, trimipramine, fluoxetine, fluvoxamine, sertraline, mianserin, moclobemide, venlafaxine, clozapine, diazepam, (es)omeprazole, lansoprazole, pantoprazole, voriconazole, brivaracetam, clobazam, lacosamide, warfarin, and labetalol.

The TraceFinder 5.1 software (Thermo Fisher Scientific, Waltham, MA, US) was used for reprocessing of full-scan HR-MS files from the TDM analyses of escitalopram. Based on a reference library (mzCloud, Thermo Fisher), comparisons of exact masses (Mw tolerance < 5 ppm) corresponding to the protonated molecular ions of small molecule drugs allowed retrospective identification of n=17 drugs in the UPLC HR-MS data files. In addition, MS/MS fragmentation patterns consistent with those of the respective compounds in the reference library were used to determine their identities. Finally, identification of specific metabolites of the respective drugs in the UPLC-HR-MS chromatograms were used and underwent the approach described above to provide supplementary evidence for correct detection of the parent compounds in the escitalopram samples.

Altogether, n=17 drugs of the initial drug list were detected in at least one patient and constitute the basis for the co-medication reported here. A small number of additional relevant drugs (clopidogrel, methylphenobarbital, and phenytoin) could not be included in the retrospective reprocessing of mass spectra from the routine analytical assay, which operates in positive ionization mode and is therefore not suitable for the reliable detection of predominantly negatively charged compounds under physiological conditions.

### Co-medication load modelling

The co-medication load was quantified as the cumulative fractional contribution of all concomitant drugs to CYP2C19-mediated metabolism. For each co-medication, the proportion of clearance attributable to CYP2C19 was extracted from the pharmacogenetic literature and expressed as a fractional value of CYP2C19 metabolism in each substrate (*fractional contribution*, Table 1) (12, 13). To predict CYP2C19 interactions, we assigned each co-medication a score based on its fractional CYP2C19 contribution as defined by (13). Scores could theoretically range from 0 (no CYP2C19 metabolism) to 1 (100% metabolism). For each participant, a *co-medication load* was then calculated by summing the fractional contribution scores of all concurrent medications; this cumulative value served as the predictor for enzyme inhibition in the first series of models detailed below. The co-medication load distribution is shown in Figure 2C.

**Figure 2.**
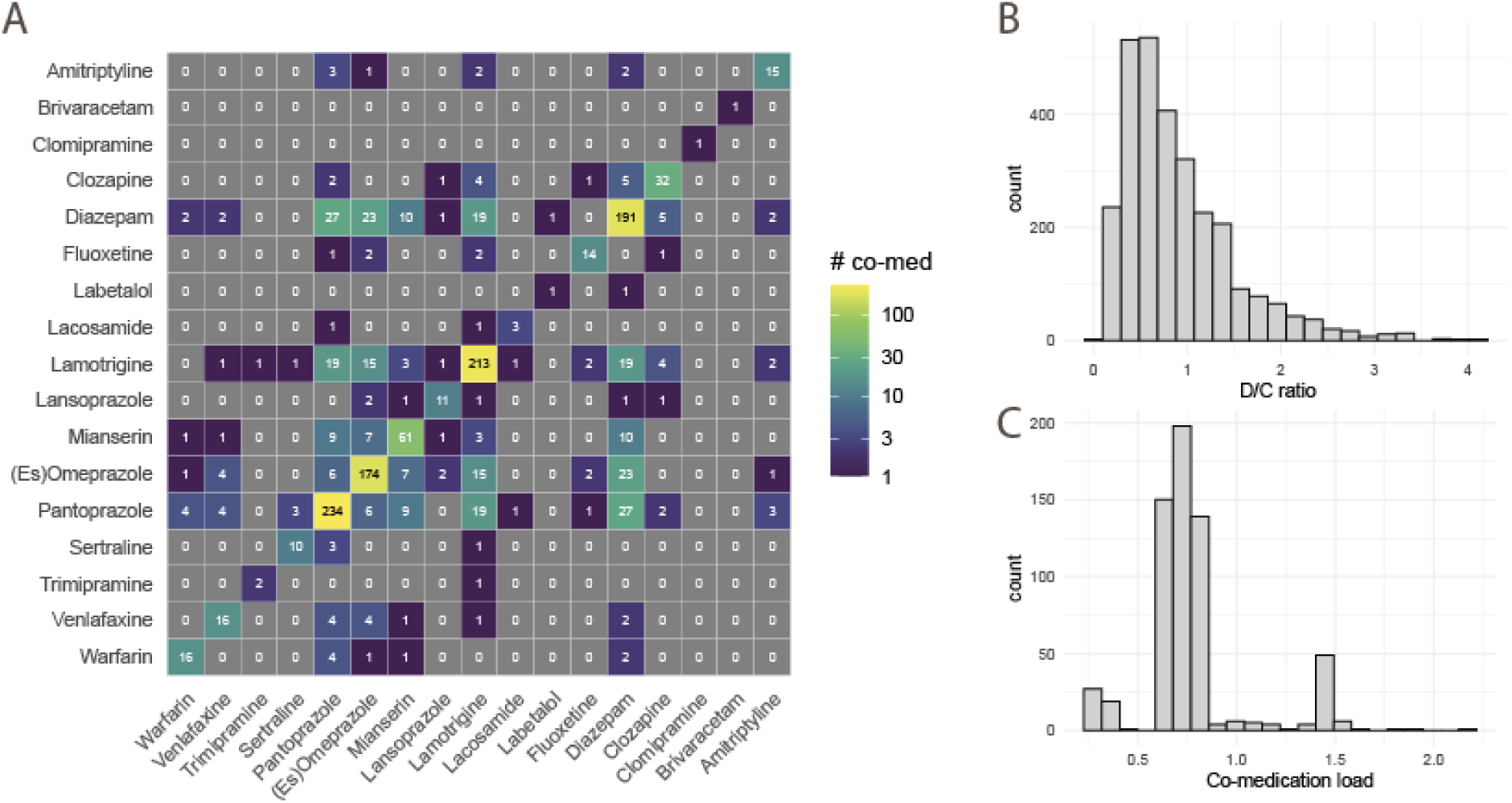
A: Number of co-medications (on the diagonal) and of co-medication combinations. # co-med: number of occurrences or co-occurrences of medication in the combinations. B: Histogram of D/C ratios in the sample. C: Histogram of co-medication load in the sample.

**Table 1:** Screened substrates and fractional contribution of CYP2C19 to their clearance from the pharmacogenetic literature (in vivo estimates)

| Medication | CYP2C19 fractional contributions | Reference |
| --- | --- | --- |
| Amitriptyline | 0.36 | Ducker et al. (13) |
| Brivaracetam | 0.30 | Stockis et al. (14) |
| Clomipramine | 0.42 | Ducker et al. (13) |
| Clozapine | 0.25 | Stingl et al. (12) |
| Diazepam | 0.65 | Ducker et al. (13) |
| Fluoxetine | 0.59 | Ducker et al. (13) |
| Labetalol | 0.66 | Chan et al. (15) |
| Lacosamide | 0.28 | Ahn et al. (16) |
| Lamotrigine | 0.00 | Mitra-Ghosh et al. (17) |
| Lansoprazole | 0.71 | Ducker et al. (13) |
| Mianserin | 0.00 | Ducker et al. (13) |
| Omeprazole | 0.80 | Ducker et al. (13) |
| Pantoprazole | 0.76 | Ducker et al. (13) |
| Sertraline | 0.61 | Ducker et al. (13) |
| Trimipramine | 0.35 | Stingl et al. (12) |
| Venlafaxine | 0.36 | Ducker et al. (13) |
| Warfarin | 0.00 | Uno et al. (18) |

### The linear phenoconversion model

The linear phenoconversion model is characterized by two assumptions: metabolic activity may be modelled as linear in activity scores appropriately chosen, and this linearity is preserved under inhibition of the enzyme by co-medication (for details and formal comparison with other models, see (6)). The expected metabolic activity is modelled as linear in the chosen activity scores, with a slope that depends on the affinity of the enzyme to the substrate (Figure 1, blue line). The intercept of this line at the PM phenotype is the metabolic activity due to other enzymes, given that no enzyme activity is present in PMs. The slope represents the effect of genetic polymorphism on the metabolism of the substrate. In this model, inhibition or induction consists in changes of the slope of this line (the inhibition is shown by light blue lines in Figure 1). The theoretically maximal possible inhibitory effect on the enzyme is a complete lack of activity, which is represented by a flat line where all phenotypes have the activity of the PM. This activity represents the contribution of other enzymes to the metabolism of the substrate.

The linearity in the model’s name is a consequence of inhibition being a proportion of initial activity (19). More precisely, if one determines the activity scores of the genotypes such that they represent the change in metabolism relative to the normal metabolizer, proportional inhibition means that the resulting linear relationship will be maintained, but with a flatter slope than in the absence of inhibition. Note, however, that inhibition by co-medication may affect enzymes other than the enzyme in question. In this case, also the metabolism in the PMs will decrease, shifting the fitted line downwards.

In a statistical model, the change in the slope of this straight line may be represented by the interaction between co-medication and the effect of the activity scores of the phenotype groups of the enzyme in question (the drug-drug-gene interaction). For co-medication inhibiting CYP2C19, we expect this interaction to be negative making a positive slope gradually approach the zero-slope flat line. The effect of co-medication at the PM estimates inhibition of other enzymes.

In the present study, this model was adapted to the TDM data by modelling phenoconversion directly from D/C ratios, instead of modelling clearance ratios. Our intent was to verify that also on this scale evidence of inhibition from co-medication was proportional to the metabolism activity in non-medicated individuals, such that the fitted slope linking phenotypic groups to CYP2C19 activity remained linear. After verifying this, we modelled the relationship between estimated changes in CYP2C19 activity due to co-medication and the fractional contribution of CYP2C19 to the metabolism of the co-medication.

### Statistical analysis

Using data from individuals without co-medication, we verified that CYP2C19 activity could be modelled in a linear relationship with activity scores from the genotype-predicted phenotypes. This preliminary analysis showed approximate linearity for activities of 0% in PMs, 50% in IMs, 100% in NMs, 115% in RMs, and 130% in UMs. This linear relationship was used as a baseline to verify that linearity held also under co-medication.

In a set of preliminary models, we established the appropriate confounder covariates (see Supplementary file 1 for details). Together with co-medication load, these covariates and the interactions with activity scores formed the predictors of the models (Table 2).

**Table 2.** Summary of terms in the model and interpretation of coefficients.

| Predictor | Interpretation of coefficient |
| --- | --- |
| Intercept | Predicted CYP2C19 activity, baseline<sup>1</sup> |
| Activity score, PM = 0 | Effect of phenotype, baseline<sup>2</sup> |
| Co-medication load | Effect of co-medication in PMs |
| Age, linear and quadratic effects | Effect of age in PMs |
| Male sex | Effect of male sex |
| Escitalopram log dose | Dose in PMs (nuisance variable) |
| Log sampling time | Log sampling time in PMs (nuisance variable) |
| Interaction activity score x co-medication load | Phenoconversion due to co-medication |
| Interaction activity score x age (linear and quadratic) | Phenoconversion analog due to age |
| Interaction activity score x log dose | Nuisance variable |
| Interaction activity score x log sampling time | Nuisance variable |

Notes. <sup>1</sup> Predicted D/C ratios in PMs, non-medicated, average age, blood sampled at 24h, escitalopram dose 10mg. <sup>2</sup> Effect of phenotype for one-unit increases in activity scores (from PM to NM) in the non-medicated of average age, blood sampled at 24h, escitalopram dose 10mg

Two sets of analyses were conducted. In the first set of analyses, we fitted three alternative models to the data with the predictors of Table 2 but using different approaches to account for the varying dispersion of residuals. They all gave qualitatively very similar predictions (see Supplementary file 2 for details on the models and results). The first was a linear model with varying residual variance as a function of the activity scores. Residual variance increased with the square of the activity scores, suggesting modelling the data in log space. Accordingly, the second model was a gamma generalized linear model (GLM) with a log link. This model fitted the data well. However, it constrains the effects of all predictors to be multiplicative — a proportional change in predicted CYP2C19 activity. Because activity scores are a strong predictor of CYP2C19 activity, this model implicitly absorbs interactions between activity scores and other predictors into the main effects, making the two unidentifiable. Since the interactions model inhibition of CYP2C19 and main effects that of other enzymes, the two mechanisms become in turn unidentifiable. To remedy this, the third model was a gamma GLM with an identity link (see ref. (20), pp. 294-295). Originally developed to model sums of squares in variance-component models, it was used here to accommodate the varying dispersion and skew of residuals while preserving the additive structure of the predictors, and with it the identifiability of main effects and interactions:

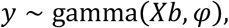

where *y* is the D/C ratio, *X* is the matrix of predictors, *b* their coefficients, and *φ* the shape parameter of the gamma distribution. This model was used in the Results section (however, details on all models are in the Supplementary file 2). Evidence for phenoconversion may be assessed by the coefficient of the interaction between activity scores and co-medication load. Evidence for non-linearity was sought by adding a quadratic term to this interaction. We also fitted interpolating linear splines after binning the observations depending on the degree of co-medication load, using general cross-validation to smooth the curves, and overlaid the estimated curves on the linear estimates of the model in figures, so as to visualize the possible departure of curves fitted to the data from the linear fit.

Models were estimated in R (The R Statistical Foundation, https://www.r-project.org/, version 4.2.2). To model varying variance, the function *gls* from the *nlme* package was used (version 3.1-160, Pinheiro and Bates (21)). Splines were fitted with the *gam* package (version 1.8-41 (22)) using a gamma GLM with identity link as in the main model. Plots were generated with the *ggplot2* package (23).

In a second set of analyses, we estimated the amount of inhibition of individual co-medications from the data themselves, instead of using the fractional contribution estimates from the pharmacogenetic literature. In the gamma GLM with identity link, the co-medication load predictor was replaced by the sum of the number of co-medications (representing the average effect of co-medication) in the main effect and in the interaction with the activity scores. Each co-medication was also modelled separately as a dummy variable (1 when present, 0 when absent), giving a predictor matrix *Z*_1_. A second matrix *Z*_2_ captured the interaction between each co-medication and the activity scores. The coefficients of these matrices estimated the deviation of individual co-medications from the average effect of adding one co-medication in the main effect and in the interaction with the activity scores. To obtain stable estimates of these coefficients, they were modelled as sampled from *t* distributions with 6 degrees of freedom:

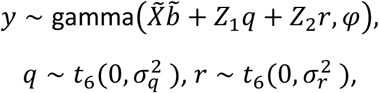

where *X̃* is the modified matrix of predictors and *b̃* their coefficients, *q* and *r* are the estimates of main deviations from average effects of individual co-medications and their interactions with the activity scores with respective variances 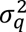 and 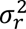. We used a Bayesian model to obtain stable estimates of these coefficients with mildly informative priors, given that some co-medications appeared only in few individuals (the estimate of the effect of these rarely detected co-medications approaches the average estimate of adding a co-medication, but with large credibility intervals; for a related model, see (12)). The *t* distribution imposes a moderate amount of shrinkage to coefficients estimated from few datapoints.

The model was fit with the freely available package *Stan* (Stan Development Team, https://mc-stan.org/) from R using the *rstan* library (version 2.32.7, Stan Development Team). Four chains were used to resample from the posterior distribution (2000 warm up and 4000 post-warm up draws per chain, giving 16000 draws for inference). To verify convergence, we used the diagnostic tools of this package. We observed no divergent transitions, effective sample sizes were considered adequate by the diagnostics, and Rhat was less than 1.005 for all estimated coefficients. Plots of coefficients and credibility intervals were drawn with the plot function of *rstan*. The code used to program and fit this model is in Supplementary file 3.

## Results

Co-medications were detected in n=817 of the n=2,852 individuals (28.6%). Figure 2A shows the count of the individual co-medications that could be identified from the spectral patterns of the reference library. Table 3 shows a break-down of the genetic phenotypes and the frequency of co-medication with one or more of the screened drugs within each phenotype group. One can see that the sample contained few individuals with more than two co-medications.

**Table 3.**
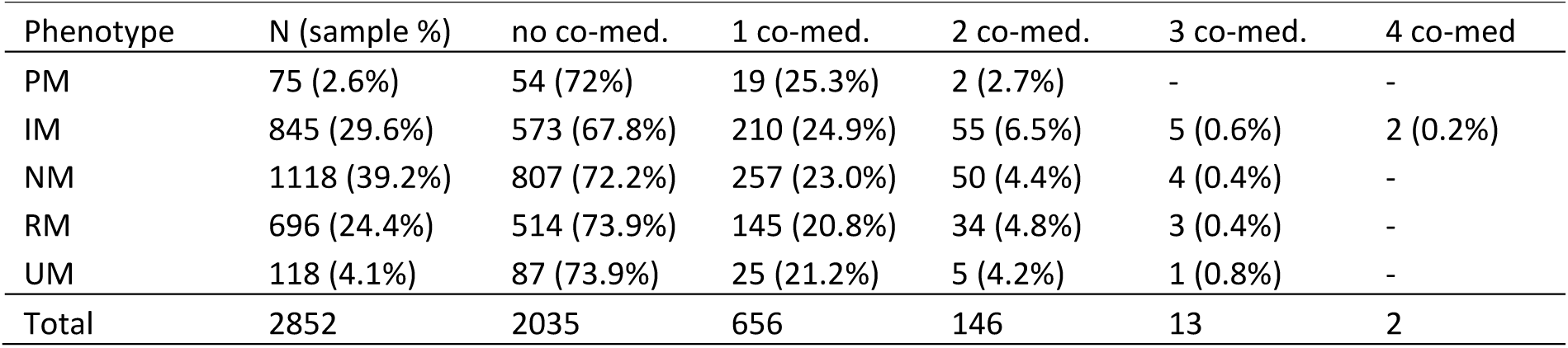
Phenotype and co-medication occurrences and frequencies.

| Phenotype | N (sample %) | no co-med. | 1 co-med. | 2 co-med. | 3 co-med. | 4 co-med |
| --- | --- | --- | --- | --- | --- | --- |
| PM | 75 (2.6%) | 54 (72%) | 19 (25.3%) | 2 (2.7%) | - | - |
| IM | 845 (29.6%) | 573 (67.8%) | 210 (24.9%) | 55 (6.5%) | 5 (0.6%) | 2 (0.2%) |
| NM | 1118 (39.2%) | 807 (72.2%) | 257 (23.0%) | 50 (4.4%) | 4 (0.4%) | - |
| RM | 696 (24.4%) | 514 (73.9%) | 145 (20.8%) | 34 (4.8%) | 3 (0.4%) | - |
| UM | 118 (4.1%) | 87 (73.9%) | 25 (21.2%) | 5 (4.2%) | 1 (0.8%) | - |
| Total | 2852 | 2035 | 656 | 146 | 13 | 2 |

Based on the conceptual framework of linear phenoconversion and the methodological approach described above (see Methods), we evaluated the effect of co-medication on CYP2C19-mediated metabolism using two complementary modelling strategies. These approaches were designed to address distinct but related questions. First, whether literature-derived information on CYP2C19 involvement of concomitant drugs can explain the observed inhibition in real-world data. Second, whether the linear phenoconversion model could independently infer the magnitude of competitive inhibition for individual co-medications directly from TDM data, and whether these data-driven estimates aligned with their known CYP2C19-dependent metabolic contribution. In both models, the reciprocal of the dose to concentration ratio of escitalopram was used as an assessment of CYP2C19 activity (Figure 2B).

These two questions were addressed by two models. In the first, we used estimates of the fractional contribution of CYP2C19 from the literature to predict the extent of inhibition of CYP2C19 metabolic capacity arising from co-medication These fractional contributions were summed across all concomitant drugs to obtain a predictor representing the overall individual co-medication load (Figure 2C). This model allowed us to assess both the presence of inhibition due to co-medication and its proportionality, that is, whether the relationship between activity scores and D/C ratios remained linear relative to the baseline linearity in non-medicated individuals.

In the second model, we estimated the magnitude of inhibition attributable to each co-medication directly from the changes in concentration to dose (TDM data), without incorporating prior information on fractional metabolic contributions from the literature. The aim of this approach was to examine the ability of the linear phenoconversion model to recover individual estimates of competitive inhibition and to relate these to the known fractional contribution of CYP2C19 to the metabolism of the respective co-medications.

### Quantitative phenoconversion from fractional contributions to the CYP2C19 metabolism of co-medication

This model found a significant effect of inhibition increasing with co-medication load (Figure 3 and Table 4). In the Figure, the data were binned into 4 groups according to the co-medication load range, and the predicted change in the linear relationship between activity scores (x axis) and CYP2C19 phenoconversion estimated by the model (y axis) was plotted for these groups. The group with load > 1.6 was too small to give indicative results (plotted with a dashed line). One can see that this relationship flattened with increasing co-medication load, as hypothesized. The inference for this progressive flattening is given by the interaction between activity score and co-medication (*p* < 0.001, Table 4).

**Figure 3.**
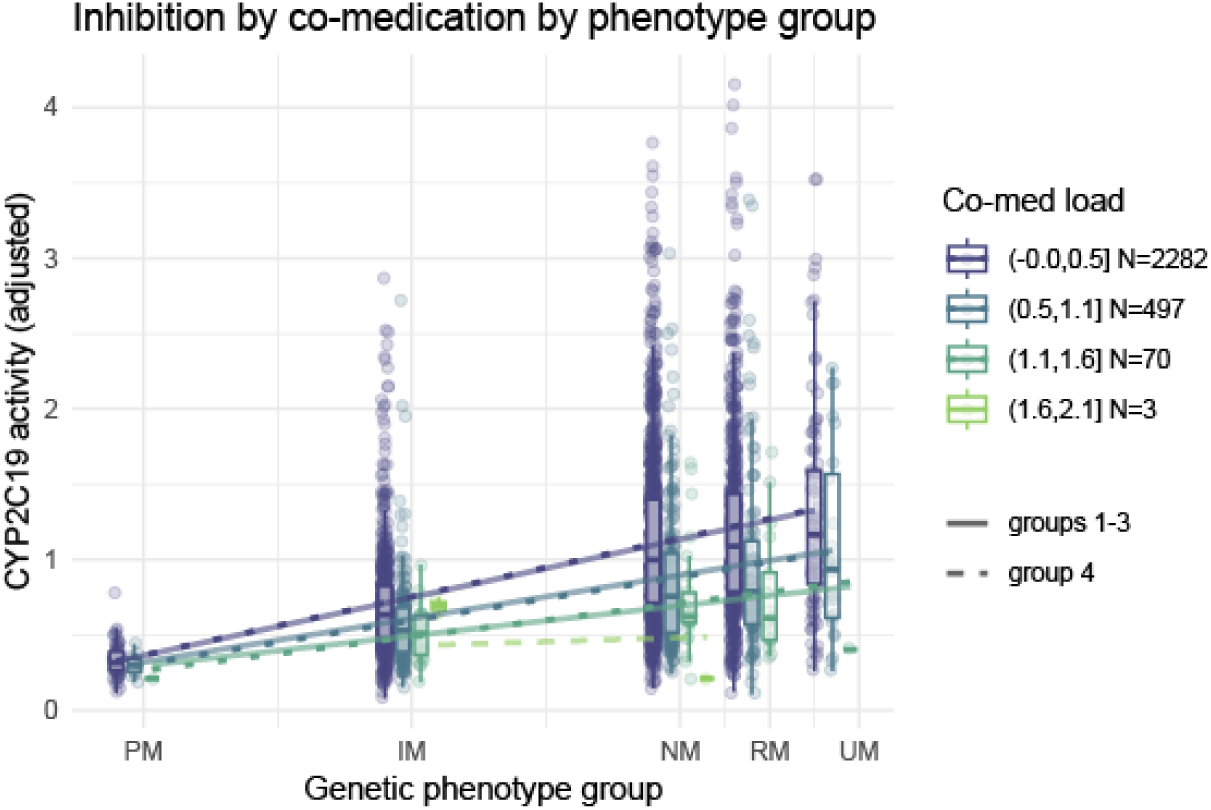
Model estimates of the interaction between co-medication load (blue to green) and activity scores (on the x axis) on D/C ratios (y axis). The box plots of the data show the approximately proportional decrease in activity in the phenotype groups with increasing co-medication load. For illustration purposes, the co-medication load was divided into four equally spaced bins (the fourth bin was too small to give indicative results). Superimposed dotted lines show a spline fit of data from each bin, regularized by cross-validation, giving essentially the same result as the linear fit of the whole model.

**Table 4.** Fit coefficients of the model of CYP2C19 activity on co-medication load.

| Coefficient | Value | Std. Err. | t-value | p-value |
| --- | --- | --- | --- | --- |
| (Intercept) | 0.32 | 0.024 | 13.40 | < 0.0001 |
| Activity score | 0.77 | 0.033 | 23.57 | < 0.0001 |
| Co-medication load (in PMs) | -0.016 | 0.039 | -0.40 | 0.69 |
| Age (linear, in PMs) | -0.033 | 0.021 | -1.57 | 0.12 |
| Age (quadratic, in PMs) | -0.0037 | 0.013 | -0.28 | 0.78 |
| Log sample time (hours, in PMs) | -0.037 | 0.053 | -0.70 | 0.48 |
| Log Escitalopram dose (in PMs) | 0.0044 | 0.032 | 0.14 | 0.89 |
| Male sex | -0.060 | 0.017 | -3.6-0 | < 0.001 |
| Activity score x co-medication load | -0.27 | 0.050 | -5.39 | < 0.0001 |
| Activity score x age (linear) | -0.048 | 0.027 | -1.80 | 0.07 |
| Activity score x age (quadratic) | -0.046 | 0.017 | -2.73 | 0.006 |
| Activity score x log sample time | 0.32 | 0.068 | 4.72 | < 0.0001 |
| Activity score x log escitalopram dose | 0.112 | 0.042 | 2.66 | 0.008 |

In this model, the relationship between activity scores and CYP2C19 activity in the non-medicated is linear by construction. To test that the relationship remained linear under co-medication, we used two approaches. In the first, we added a quadratic activity-score term to the activity-score × co-medication load interaction. This term, however, was not significant (*t* = −0.42, *p* = 0.68; not shown in Table 4, see Supplementary file 2). In the second, we fitted the adjusted CYP2C19 activity to the activity scores separately in the phenotype groups using an interpolating spline regularized by cross-validation. This fit will give a curve in the co-medication groups if the relationship between activity scores and CYP2C19 activity deviates from linearity. The fit is shown as superimposed dotted lines. As shown in Figure 3, these fits could not be distinguished from the linear fits, consistent with the lack of significant quadratic effects of the previous model. We conclude that the relationship showing the increase in CYP2C19 activity when moving from the PM to the UM groups remained linear and progressively flattened due to the increasing inhibition, as in the linear phenoconversion model (Figure 1). This conclusion, however, is limited to co-medication loads up to about 1.6, as the data did not contain enough information on heavier co-medication loads. For the same reason, we could not model the saturation of the inhibition that inevitably arises when the metabolic activity reaches that of the PMs.

Table 4 quantifies estimated effects of all predictors in the model. In observations without known co-medication, assuming mean age, a sample time of 24 h, and an escitalopram dose of 10 mg, the estimated activity score coefficient was 0.77 kL/day. This coefficient represents the difference in CYP2C19 activity between normal metabolizers (NMs) and poor metabolizers (PMs). The interaction with co-medication load represents phenoconversion. One unit of fractional CYP2C19 contribution in the co-medication reduced this coefficient by 0.27 to 0.50 kL/day, i.e. to about two thirds of the original NM–PM difference.

To give a term of comparison, we show in Figure 4 the progressive inhibition due to age estimated by the same model (linear and quadratic effects, *F*(2, 2839) = 22.0, *p* < 0.001). The figure shows that the relationship between activity scores and age remained linear with increasing age, but age started to affect metabolic capacity only from about 60 years. However, age was also a significant inhibitor of metabolic capacity in PMs (*F*(2, 2839) = 3.1, *p* < 0.047). For this reason, the inhibitory effects of age cannot be modelled within the range of the genetic polymorphism, as they are not bounded below by the metabolism in PMs.

**Figure 4.**
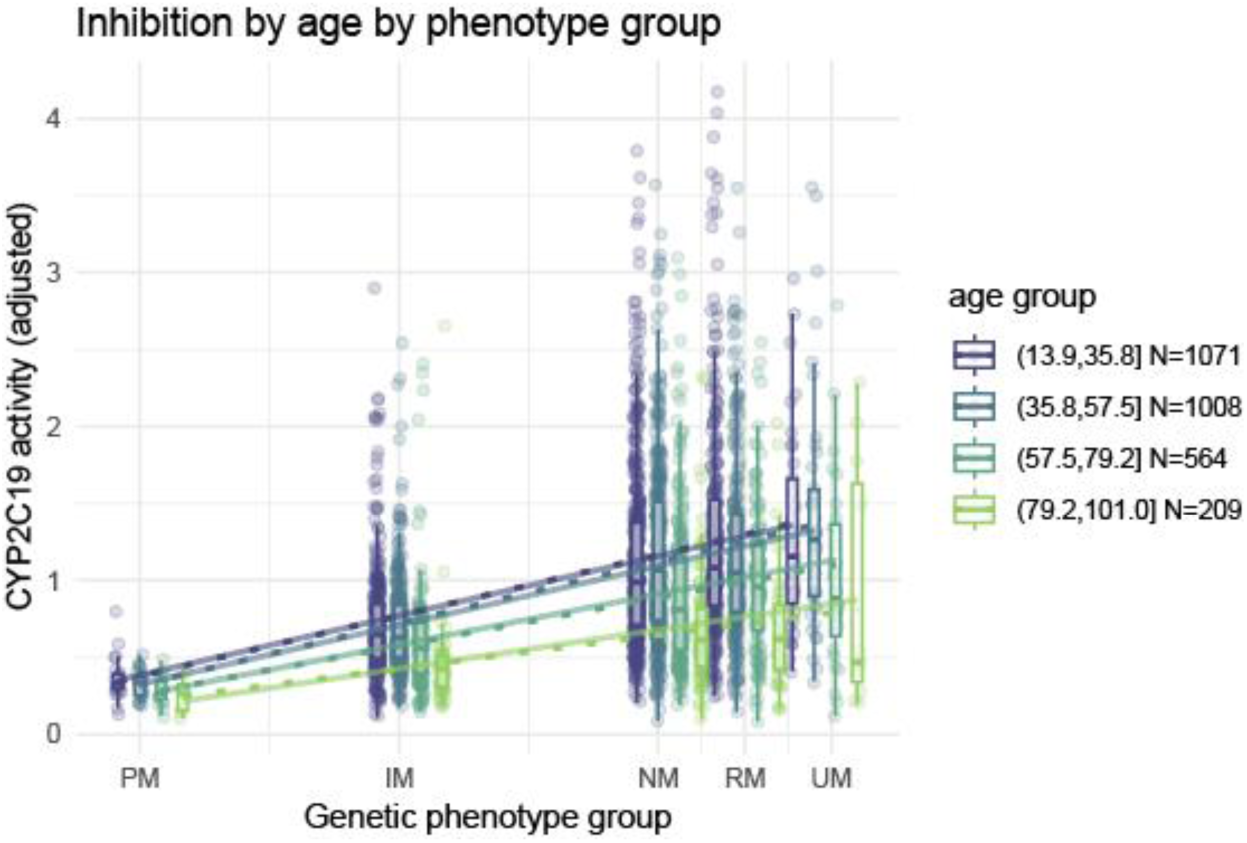
The same plot as in the previous figure, this time grouped by age to show the interaction between this predictor and the activity scores. The same linear relationship in all age groups is also held here.

### Inhibition by individual co-medication

After having established a statistical model for the linear phenoconversion, we turned to the question of whether this model can extract information on inhibition by individual co-medication from the TDM data. If this is the case, we should recover quantities correlated with the fractional activities of CYP2C19 in the metabolism of the co-medication that we used as predictors in the previous section.

To this end, we adopted a Bayesian approach to model the amount of CYP2C19 phenoconversion in individual co-medications. Crucially, information on the fractional contribution of CYP2C19 to the metabolism of individual medications (Table 1) was not used in this model. Instead, the model included dummy variables-predictors representing the presence of each co-medication, one per co-medication, to allow estimating the inhibiting effect of each co-medication separately from the data themselves. As in the previous model, phenoconversion was modelled by the interaction of these individual co-medication predictors with the activity score of the genetic phenotype, while the main effects of co-medications variables represented effects on CYP2C19 activity in the PM (inhibition of enzymes other than CYP2C19). This model required a Bayesian approach to stabilize the fit, since for some medications there were very few or just one observation (in this case the coefficients show the mean effect of adding a medication with a large credibility interval).

In the plot below (Figure 5), the estimated interaction of individual co-medication with the activity scores is shown (inhibition of CYP2C19). None of the effects on CYP2C19 activity in PMs (which could be the case if other enzymes than CYP2C19 would be affected by inhibition as well) was individually significant.

**Figure 5.**
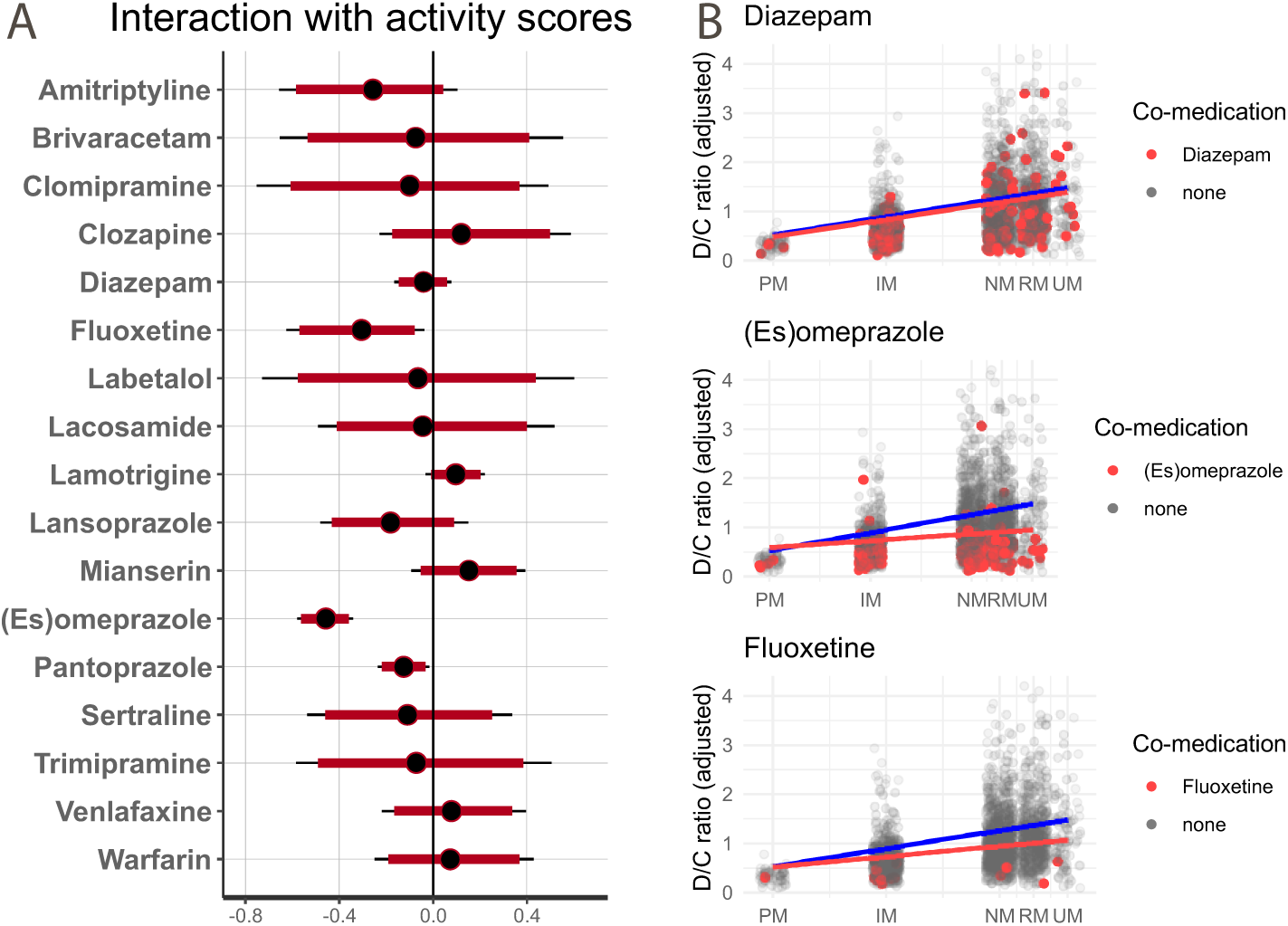
A: Estimates of inhibition of CYP2C19 of individual medication for the metabolism of escitalopram (interaction of individual co-medications and activity scores, changes in kL/day). The thick intervals are effects at trend level, two-tailed (95% intervals). The thin lines are significant at 5%, two-tailed. The black dots are the median estimates. A negative effect corresponds to inhibition. B: Examples of estimated fits for individual co-medications (red). The blue line corresponds to the predicted CYP2C19 activity in non-medicated individuals.

As can be seen from the figure, significant CYP2C19 inhibition was detected for fluoxetine, (es)omeprazole and pantoprazole (Table 5).

**Table 5.** Estimates of individual co-medication inhibitory effects on CYP2C19 activity, on other metabolic pathways, and relative 95% credibility intervals.

| Co-medication | CYP2C19 | 2.5% | 97.5% | other | 2.5% | 97.5% |
| --- | --- | --- | --- | --- | --- | --- |
| Amitriptyline | -0.263 | -0.656 | 0.103 | 0.129 | -0.130 | 0.456 |
| Brivaracetam | -0.070 | -0.654 | 0.554 | -0.017 | -0.386 | 0.356 |
| Clomipramine | -0.107 | -0.752 | 0.492 | -0.006 | -0.380 | 0.381 |
| Clozapine | 0.136 | -0.229 | 0.586 | -0.075 | -0.363 | 0.205 |
| Diazepam | -0.041 | -0.166 | 0.078 | -0.037 | -0.120 | 0.058 |
| Fluoxetine | -0.312 | -0.626 | -0.036 | -0.008 | -0.194 | 0.238 |
| Labetalol | -0.066 | -0.728 | 0.602 | -0.074 | -0.442 | 0.294 |
| Lacosamide | -0.028 | -0.491 | 0.518 | -0.046 | -0.393 | 0.273 |
| Lamotrigine | 0.097 | -0.031 | 0.220 | -0.048 | -0.129 | 0.046 |
| Lansoprazole | -0.178 | -0.481 | 0.150 | 0.034 | -0.199 | 0.284 |
| Mianserin | 0.153 | -0.092 | 0.392 | -0.064 | -0.211 | 0.113 |
| (Es)omeprazole | -0.459 | -0.580 | -0.340 | 0.065 | -0.033 | 0.172 |
| Pantoprazole | -0.125 | -0.237 | -0.016 | -0.019 | -0.101 | 0.069 |
| Sertraline | -0.064 | -0.537 | 0.336 | 0.030 | -0.249 | 0.333 |
| Trimipramine | -0.064 | -0.584 | 0.506 | -0.017 | -0.376 | 0.340 |
| Venlafaxine | 0.082 | -0.220 | 0.395 | -0.051 | -0.297 | 0.225 |
| Warfarin | 0.079 | -0.249 | 0.428 | -0.056 | -0.336 | -0.061 |

We then verified the association between these estimates of phenoconversion and the pharmacogenetic-derived estimates of the CYP2C19 fractional contribution of the co-medication from the literature (Table 1). We found that the literature fractional contributions of CYP2C19 predicted the inhibition estimated from the TDM data (*t*(15) = −4.41, *p* < 0.001), accounting for about 53% of its variance.

As shown in Figure 6, most of the co-medications inhibited escitalopram metabolism as predicted by the fractional contribution of CYP2C19 to their own metabolism. The exceptions were fluoxetine and (es)omeprazole, where metabolism of escitalopram was lower than predicted from their fractional contributions. According to the model, a 0.5-unit increase in the fractional contribution corresponded to a flattening of the activity-score slope of about 0.23 kL/day — roughly a 30% reduction in the CYP2C19-mediated metabolic capacity of escitalopram relative to its uninhibited value — that is, a phenoconversion of the genetic phenotype toward lower effective activity.

**Figure 6.**
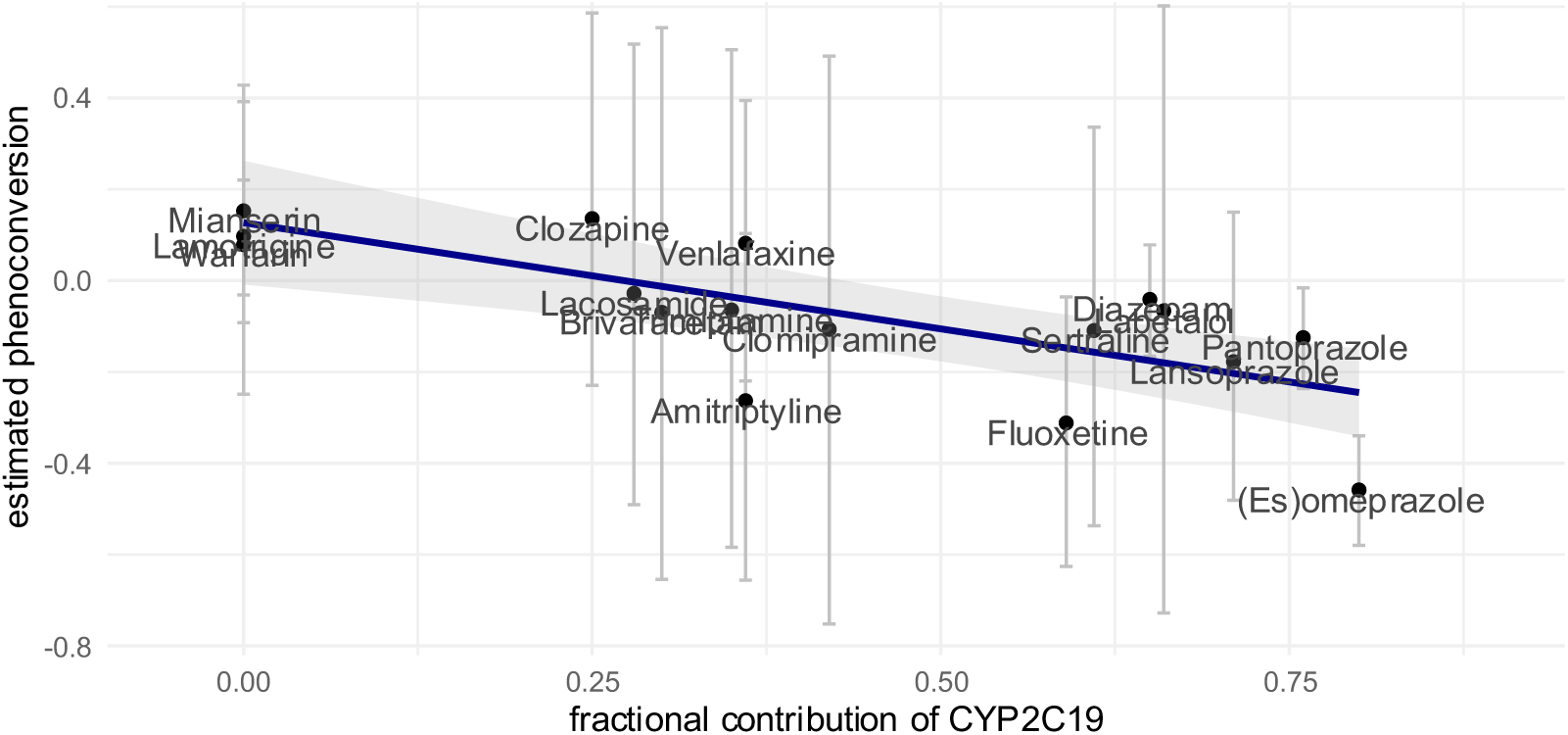
Scatterplot of the estimated phenoconversion coefficients of individual co-medications vs. estimates of the fractional contribution of CYP2C19 to their metabolism from the pharmacogenetic literature. The vertical bars are the 95% credibility intervals of the estimated inhibition from the co-medication from the model of the previous figure.

In the plot, we see that substrates for which we had few observations have large credibility intervals (vertical bars) and, except for labetalol, cluster around the middle of the regression line, where the fitted inhibition was shrunk toward zero by the model. Therefore, the regression estimate is heavily influenced by the substrates with no affinity for CYP2C19 and the few substrates with strong inhibition.

To verify the sensitivity of the inference to these influences, we removed from the dataset fluoxetine and omeprazole (with high inhibition estimates), and the substrates with fractional contributions of zero (at the opposite extreme of the inhibition spectrum). The correlation remained significant, even if less than in the first model (*t*(10) = -3.76, *p* = 0.004).

## Discussion

Pharmacogenetics has long been recognized as a valuable tool in psychiatry, with well-established dosing recommendations and evidence-based guidelines, such as those provided by the Clinical Pharmacogenetics Implementation Consortium (CPIC), for both antidepressants and antipsychotics (1, 24, 25). However, the meaningful integration of pharmacogenetic information into individualized treatment selection and dose optimization requires simultaneous consideration of additional key determinants of drug response. Among these, co-medication represents a critical yet often underappreciated factor due to its potential to modulate drug metabolism (2). In the present study, we developed a quantitative model that enables estimation of the impact of concomitant medications on CYP2C19-mediated metabolism. Our findings demonstrate that knowledge of the fractional contribution of CYP2C19 to the metabolism of a given substrate is sufficient to predict the extent of phenoconversion, as the magnitude of this effect directly reflects the enzyme’s relative involvement. Consequently, classical pharmacogenetic dose recommendations based on clearance ratios (e.g., (25–27)) prove highly informative for approximating the degree of phenoconversion resulting from substrate inhibition. Furthermore, in the context of polypharmacy, we show that a simple additive approach can be applied to estimate the cumulative impact on CYP2C19 activity, thereby facilitating more accurate, integrative predictions of drug metabolism in complex clinical scenarios.

The first aim of this study was to test whether phenoconversion of CYP2C19 activity by co-medication can be adequately described by a linear phenoconversion model, in which competitive inhibition leads to a flattening of the relationship between pharmacogenetic phenotype groups and metabolic activity, as originally proposed by Ohno et al. (2007) and formalized by Borges et al. (2010) and Viviani and Stingl (2025) for clearance (6, 7, 19).

Using literature-derived estimates of fractional CYP2C19 contribution of each detected co-medication, we were able to quantitatively model the extent of phenoconversion as a function of the summed metabolic burden imposed by concomitant drugs. Our results confirm earlier findings by Borges et al. (2010) that competitive inhibition preserves the linear relationship between activity score and enzyme activity, while reducing its slope relative to the non-medicated baseline (7). Thus, the fundamental assumption of linear phenoconversion was confirmed in a large real-world TDM sample comprising genotyped patients treated with the CYP2C19 substrate escitalopram.

Importantly, the use of summed fractional contribution values allowed phenoconversion to be assessed quantitatively at the individual co-medication level, even in the presence of more than one concomitant drug. This approach goes beyond categorical descriptions of drug–drug interactions and provides a mechanistically grounded representation of co-medication effects on CYP2C19 activity.

Quantitatively, the magnitude of phenoconversion observed in this study was modest when compared with the effect of genetic loss of function. Co-medication corresponding to a summed fractional CYP2C19 contribution of 1.0—equivalent to a single 100% CYP2C19 substrate or two substrates each metabolized 50% via CYP2C19—reduced the activity score coefficient by approximately one third. Therefore, even a substantial CYP2C19 substrate burden leads to moderate phenoconversion rather than a complete shift toward the PM phenotype. This quantitative contrast illustrates that inhibition by co-medication attenuates, but does not necessarily abolish, genetically determined differences in CYP2C19 activity, consistent with the assumptions of the linear phenoconversion model.

A second question of interest was whether co-medication affects escitalopram metabolism in CYP2C19 PMs. As hypothesized previously (2), inhibition of alternative elimination pathways could reduce metabolism even in the apparent absence of functional CYP2C19 activity, captured by the model as the main effect of medication in PMs. However, in our data these effects failed to reach significance. This finding supports the core concept of phenoconversion: inhibitory effects scale with the amount of functional enzyme present.

### Age effects and their distinction from phenoconversion

In contrast to co-medication effects, age had a clear and independent impact on CYP2C19 activity, including in PMs (Fig. 4). This is consistent with age-related reductions in hepatic extraction, renal elimination and body composition influencing D/C ratios, and other pharmacokinetic factors which affect drug metabolism independently of CYP2C19 genotype.

The age effect remained visible even in CYP2C19 PMs, indicating a mechanism distinct from CYP2C19 phenoconversion. Nevertheless, the age effect still interacted with metabolic capacity, leading to a linear downshift in CYP2C19 activity with a reduced slope of the activity-score relationship. Analysis of different age groups showed that CYP2C19 activity reductions occurred predominantly in older individuals (above approximately 60 years of age), justifying the inclusion of age as a quadratic term in the model.

#### Residual variability and model limitations

Although phenoconversion by co-medication was statistically significant, it explained only a limited proportion of the overall variability in escitalopram D/C ratios. Substantial residual variability remained even among subjects with detected co-medication, and this variability was comparable to that observed in the overall TDM sample.

The influence of co-medication emerges primarily through interaction with the genetic phenotype, such that its effect increases with the amount of functional enzyme available. However, numerous observations deviated from predicted CYP2C19 activity estimates, most prominently in the highly active phenotypes, where prediction uncertainty increased. This pattern suggests the presence of additional, unmeasured modifiers of CYP2C19 activity acting through similar mechanisms, i.e. unknown co-medication acting on the available functional protein.

Due to the cross-sectional design of the TDM study data, we did not have paired observations from the same individuals under conditions with and without co-medication, which limits within-subject attribution of observed changes in CYP2C19 activity to co-medication effects. Consequently, phenoconversion estimates rely on between-subject comparisons across pharmacogenetic strata rather than longitudinal observations within individuals.

Co-medication was detected in approximately 29% of escitalopram TDM-samples, most frequently involving one single additional drug. Although this does not represent the assumed prevalence of polypharmacy in the general population—particularly compared to elderly cohorts—it allowed investigation of phenoconversion in a large, well-characterized sample with available CYP2C19 genotypes. The psychiatric patient population studied here likely represents a population in which combination therapy is common, even in younger individuals.

However, while the Orbitrap-based screening enabled detection of a substantial proportion of co-medications directly from plasma samples, it cannot be assumed that all concomitant drugs influencing CYP2C19 activity were captured, as a comprehensive scan of all possible co-mediation was not feasible. Such unobserved co-medication may partly account for the residual variability in this activity, particularly in individuals with high predicted metabolic capacity. In non-medicated PMs, it may have led to underestimating CYP2C19 activity due to undetected co-medication. The failure to demonstrate inhibition of enzymes other than CYP2C19 at levels of statistical significance may be due to this limitation.

Interestingly, variance within the PM group was comparatively low in absolute terms. This suggests that sources of variability beyond functional enzyme abundance—such as measurement error or stochastic influences—have limited impact when CYP2C19 activity is absent. Residual variability in other phenotype groups may partly reflect rare genetic variants (28), regulatory mechanisms, or epigenetic effects, as well as real-world influences such as variable absorption, hepatic blood flow, dietary factors, or endogenous inhibitors. In addition, TDM-based estimation of CYP2C19 activity from a single concentration–dose ratio is inherently less precise than full pharmacokinetic profiling.

#### Medication/substrate specific inhibition

##### Substance-specific CYP2C19 inhibition findings from the second model

The analysis of substance-specific effects largely confirmed a proportional relationship between the fractional contribution of co-medications to CYP2C19-mediated metabolism and the estimated interaction term derived from the phenoconversion model. For most co-medications included in the present dataset, the observed degree of inhibition closely followed the expected magnitude based on the reported fractional contribution of CYP2C19 to their metabolism, supporting the assumption that competitive inhibition by CYP2C19 substrates can be adequately captured by a linear phenoconversion framework.

Deviations from a proportional relationship between fractional CYP2C19 contribution and the interaction term would be expected for inhibitors that do not act solely as competitive substrates of CYP2C19. For most co-medications included in the present dataset, no such deviations were observed. However, for (es)omeprazole—and to a lesser extent fluoxetine—systematic deviations from the expected proportionality were evident. (Es)omeprazole is both a substrate and a mechanism-based (time-dependent) inhibitor of CYP2C19, whereas fluoxetine is a moderate CYP2C19 inhibitor with predominantly competitive inhibition and weak metabolism-dependent effects mediated by its active metabolite norfluoxetine. In contrast to purely competitive inhibition, mechanism-based inhibition is expected to result in stronger and more sustained effects, consistent with the patterns observed for (es)omeprazole and, to a lesser extent, fluoxetine in Figure 5 (29, 30).

Importantly, apart from these well-characterized cases, no systematic departures from the expected proportional relationship were identified. This high concordance between literature-based fractional contribution estimates and data-driven inhibition terms support the validity of the fractional contribution approach as a quantitative predictor of phenoconversion in real-world settings.

##### Medications without expected CYP2C19 effects

Lamotrigine was among the most frequent co-medications. Although interaction warnings exist for lamotrigine and SSRIs, these are largely pharmacodynamic. Lamotrigine is metabolized predominantly by UGT enzymes and active transport mechanisms, and its intrinsic CYP2C19 fractional contribution was therefore set to zero (17). Consistently, no phenoconversion effect was observed. Mianserin and warfarin, which are not metabolized by CYP2C19 (13, 18), also showed no phenoconversion effects.

##### Antidepressants and anxiolytics

Diazepam and sertraline showed little or no CYP2C19 inhibition effect, despite reported CYP2C19 involvement. This may reflect differences in relative affinity between escitalopram and these substrates, or inversion of perpetrator–victim roles. Notably, diazepam is widely considered a CYP2C19 victim drug rather than a perpetrator, and no CYP2C19 inhibition warnings are issued for diazepam affecting other substrates (31). Sertraline’s lack of effect may further relate to allele-specific inhibition patterns, with weaker inhibition of the CYP2C19*1 allele compared to other variants (32). Tricyclic antidepressants showed consistent, moderate phenoconversion in line with their fractional contribution values (amitriptyline strongest, trimipramine and clomipramine less). Venlafaxine and lacosamide showed minimal phenoconversion, but case numbers were limited.

## Conclusion

Taken together, these findings establish linear phenoconversion as a robust and mechanistically grounded framework for integrating pharmacogenetic variability with co-medication effects on CYP2C19 activity. From a clinical psychiatry perspective, this approach directly addresses a central challenge in psychopharmacology: the reliable estimation of drug exposure under conditions of polypharmacy. By quantifying inhibitory burden using fractional contribution rather than categorical interaction labels, phenoconversion can be operationalized in a manner that reflects real-world prescribing complexity, thereby enabling more accurate prediction of individual metabolic capacity.

While escitalopram served as the probe and victim drug in this study, the principles described here should be applicable to other CYP2C19 substrates. By complementing drug-interaction labels with quantitative estimates of inhibition, this approach offers a route toward data-driven drug–drug–gene interaction (DDGI) assessment within clinical decision support systems. Ultimately, such integration holds promise for advancing precision dosing and improving treatment outcomes in psychiatric populations characterized by multimorbidity, comedication, and substantial inter-individual variability in drug response.

## Data Availability

Inquiries about data used in this article should be addressed to Dr. Espen Molden, Center for Psychopharmacology, Diakonhjemmet Hospital, Oslo, Norway

https://git.uibk.ac.at/c7201031/ztr2503.git

## Software availability

The markdown files that include the code used in this work (including the code generating the supplementary material files and all models used in the main text) are available at the git repository https://git.uibk.ac.at/c7201031/ztr2503.git.

## Acknowledgments

The authors declare no competing interest.

Julia Stingl was supported by ERA PerMed grant (project ArtiPro) of the German Ministry of Education and Research (BMBF, grant number: 01KU2212). Kristine Hole was supported by ERA PerMed grant (project ArtiPro) of the Norwegian Research Council (grant number 94035). Roberto Viviani was supported by ERA-PERMED grant (project ArtiPro) of the FWF Austrian Science Fund (grant number I 5903) [Grant-DOI:10.55776/I5903]. ERA PerMed is supported by funding from the European Union’s Horizon 2020 research and innovation programme under grant agreement No. 779282. For open access purposes, the authors have applied a CC BY public copyright license to any manuscript version arising from this submission.

## Notes

### Competing Interest Statement

The authors have declared no competing interest.

### Author Declarations

Regional Ethical Committee (# 482562) and the investigational research board of the Diakonhjemmet Hospital in Oslo

