## Supplemental file 1 for "Pharmacogenetic phenoconversion modeling of drug–drug–gene interactions on CYP2C19 activity: effects of comedication by genotype on escitalopram concentrations"

### Supplementary results 1: Data exploration and description

Roberto Viviani, Institute of Psychology, University of Innsbruck; Psychiatry and Psychotherapy III, University of Ulm

2025-04-04

This dataset provides information on the metabolism of escitalopram by CYP2C19 while considering common co-medications, some of which may inhibit CYP2C19 and lead to phenoconversion of the CYP2C19 genetic phenotype. The effect of co-medication is expected to be detectable in the plasma levels of escitalopram in interaction with the CYP2C19 phenotype. See the main text for the methods used to detect co-medication in the sample. In previous work with this dataset, Hole et al., (submitted) could show that statins had an effect on the CYP2C19-mediated metabolism of escitalopram.

Here, we provide a preliminary exploration of this dataset. We used the dose to concentration ratio (D/C ratio) adjusted for the log of the time of blood sampling as a measurement of the metabolism of escitalopram. This quantity is expressed in kL (kiloliters) and represents the apparent volume of distribution at the time the sampling was taken. Our working hypothesis is that, at a fixed sampling time (or equivalently after adjusting for sampling times), this quantity is a proxy for clearance values, such that modelling approaches developed for clearance may be applied here too.

We start the analysis by exploring the distribution of the D/C ratio and the relationship with possible predictors. We will then model causal interactions with a DAG to establish the necessity of including covariates. We will then examine the residuals of a preliminary model to identify observations that deviates from expectations (suspect cases of non-adherence). Finally, we will provide a description of the cleaned-up sample.

#### Preliminary exploration

The histogram of the concentration to dose ratios shows that there was an individual with an extreme outlier value (a NM with no co-medication). In the analyses below, we will prudentially exclude this individual.

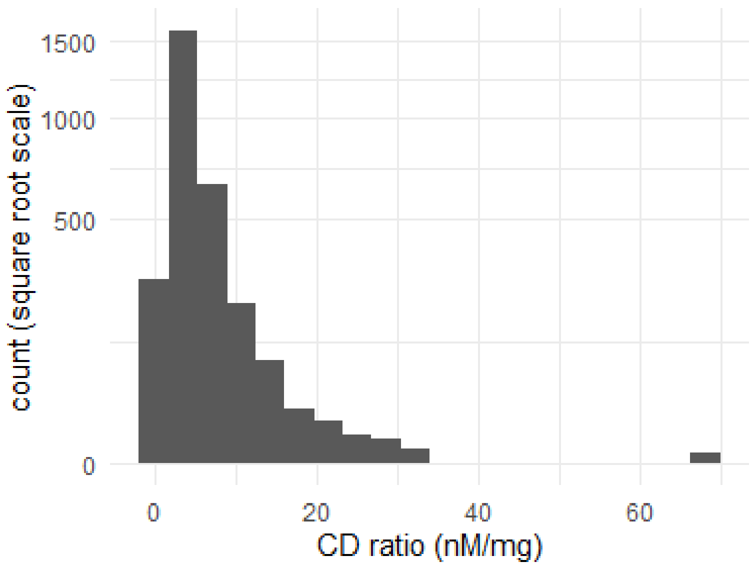


#### D/C ratios vs sampling time

The time in which plasma concentration of escitalopram was assessed by taking blood samples varied considerably in the sample.

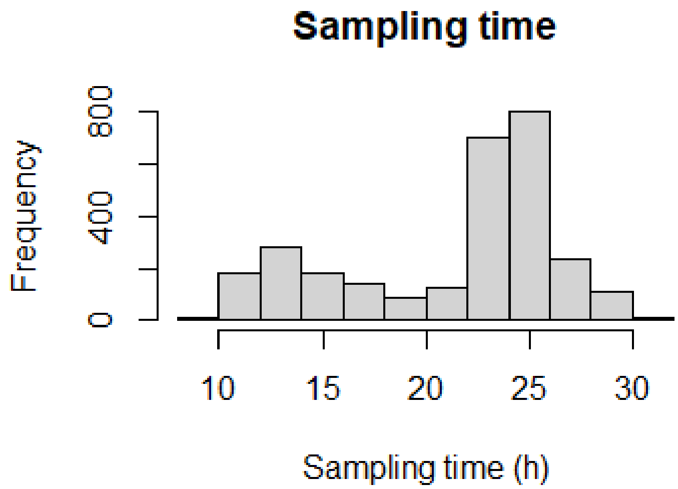


Below, the relationship between the D/C ratio and the log of time of sampling is shown.

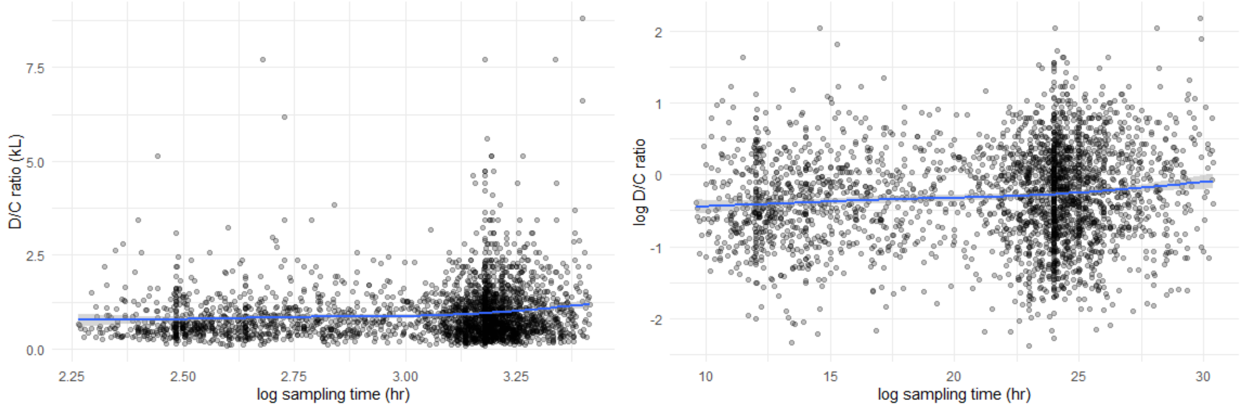


The relationship between log sampling time and D/C ratio is approximately linear, with only a slight increase at sampling times over 25 hrs, irrespective of whether D/C ratio is log-transformed or not. We will accept the linear approximation in the models that follow.

#### D/C ratio vs dose

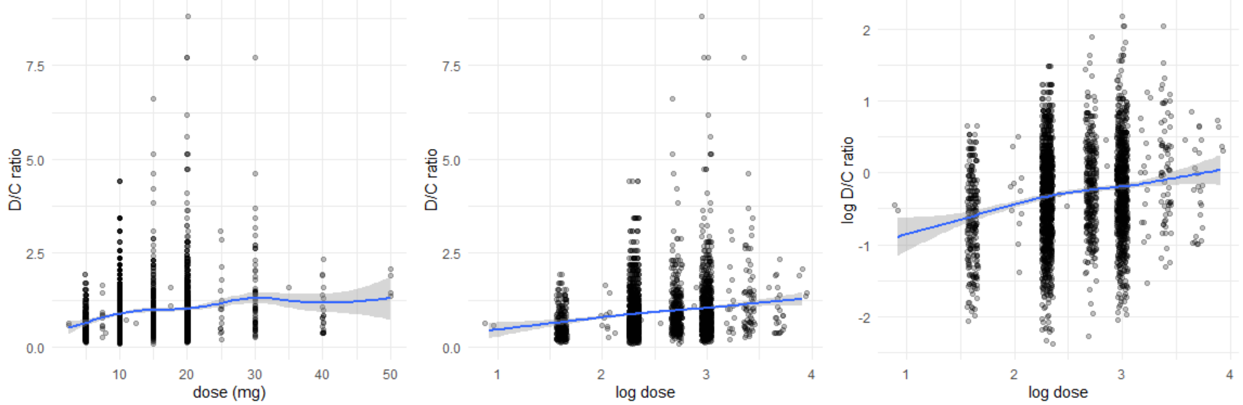


The relationship between D/C ratio and dose is best characterized by taking the log of dose. In this case, the relationship is approximately linear. The effectiveness of the log transformation here may be understood as adjusting for the measured escitalopram concentration, which is in the numerator in the D/C ratio used as the output variable. Therefore, in the models that follow we adopted log dose as a candidate confounder.

In all models examined below the quadratic effect of age was strongly significant, as is apparent from the plots below.

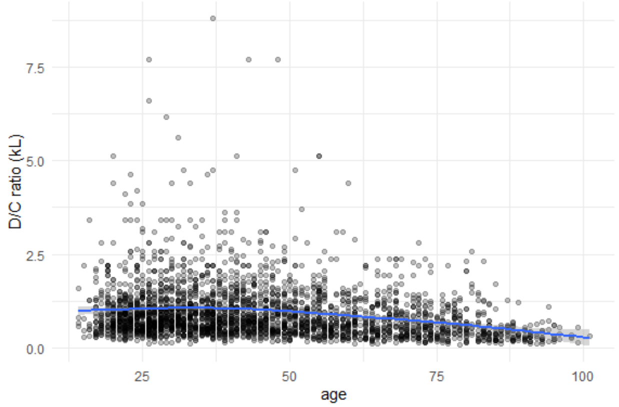


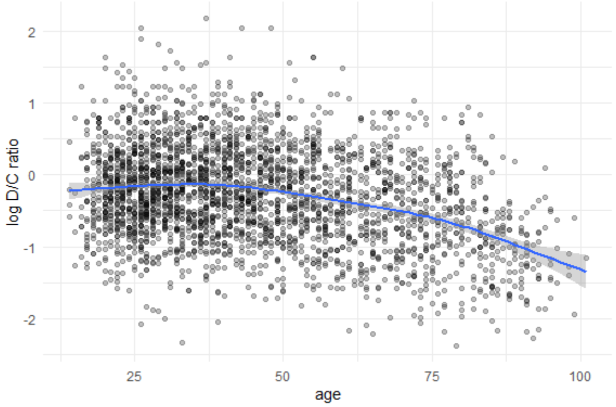


#### Co-medication load and number of medications

The hypothesis of the study was that the *co-medication load*, a quantity derived by summing the fractional contribution of CYP2C19 to the metabolism of the co-medications concomitantly taken by the individual, is a predictor of the inhibition of escitalopram metabolism (competitive inhibition). This quantity is also related to the number of co-medications taken:

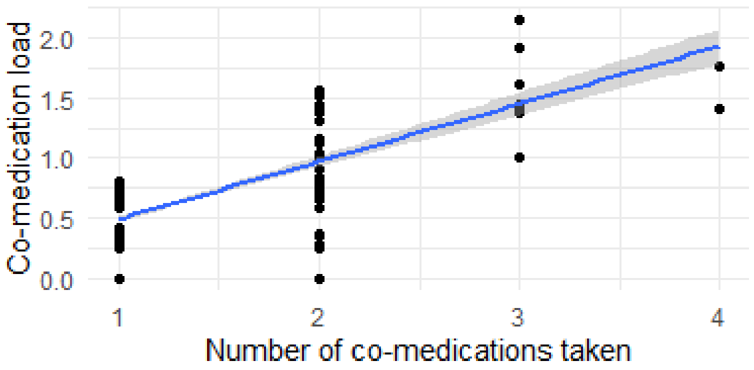


In the analyses below, we will use number of co-medications to examine the causal model to apply to the data, as the number of co-medication is interpretable as being caused by other variables (for example, age will increase the number of medications taken). However, in the main modelling sections, we will use co-medication load as a predictor.

### Causal model and distribution of residuals

In this section, we conduct a preliminary data screening to identify potential confounders. To start, we will use a simple linear regression model. In a causal framework, a confounder must be associated with both the exposures (CYP2C19 phenotype and co-medication) and the outcome (D/C ratio). We examine these associations independently to build a Directed Acyclic Graph (DAG), ensuring that we identify variables that could bias our estimates. More generally, we will be also interested in variables that are associated with the outcome and may be added as covariates to the model without biasing estimates according to the DAG model. Furthermore, we evaluate the distribution of residuals to ensure our model assumptions—particularly regarding the functional form of D/C ratios—are met.

#### Confounders by association with the outcome variable

It appears that all variables in the dataset are associated with the D/C ratio, including log sampling time, log dose, age and its quadratic term, and sex. As one might expect, also co-medication (summarized here by the TotalMeds variable, the number of co-medications, as an index of potential inhibition of CYP2C19 activity) was associated with the D/C ratio of escitalopram.

```
MODEL INFO:
Observations: 2872
Dependent Variable: DC_ratio
Type: OLS linear regression

MODEL FIT:
F(6,2865) = 54.12, p = 0.00
R² = 0.10
Adj. R² = 0.10

Standard errors: OLS
-----------------------------------------------------------
                                Est.   S.E.   t val.      p
---------------------------- ------- ------ -------- ------
(Intercept)                    -0.60   0.16    -3.71   0.00
log(Sampling_time_ESC)          0.36   0.05     7.84   0.00
log(ESC_dose)                   0.20   0.03     6.80   0.00
SexM                            0.10   0.03     3.79   0.00
Age.z                          -0.10   0.02    -6.26   0.00
I(Age.z^2)                     -0.06   0.01    -4.46   0.00
TotalMeds                      -0.13   0.02    -5.88   0.00
-----------------------------------------------------------
```

A look at the residuals of these first preliminary analyses shows a strong heteroscedasticity. They form a funnel expanding from the low to the high predicted values, and are also higher on the upside (while the D/C ratio cannot be negative, it can be very high).

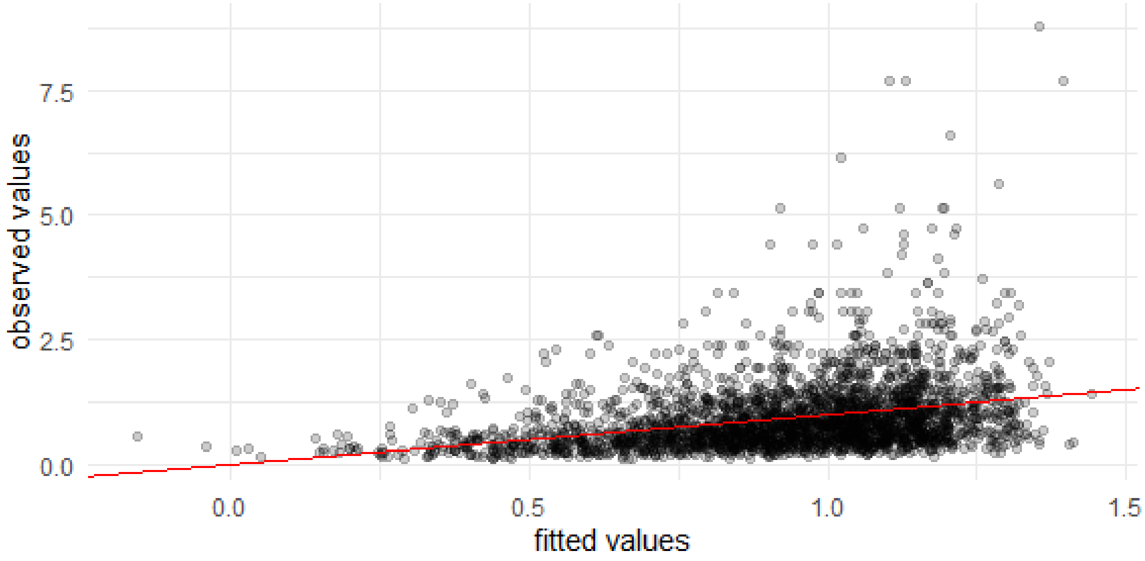


The funnel pattern of residuals is what one would expect if effects on the empirical phenotypes were fractional changes of a baseline D/C ratio, i.e. multiplicative effects (which suggest a log-transform of the outcome for the model). Modelling this variance will be a central issue in the analysis that follows, including the consequences of log-transforming the outcome. For now, we verify that the associations concerning possible confounders are present after log-transforming D/C ratio values. As one may expect from their magnitude, these effects were not influenced appreciably by this transformation. Hence, the same adjustment model remains appropriate irrespective of the transformation of the outcome variable.

```
MODEL INFO:
Observations: 2872
Dependent Variable: log(DC_ratio)
Type: OLS linear regression

MODEL FIT:
F(6,2865) = 96.99, p = 0.00
R² = 0.17
Adj. R² = 0.17

Standard errors: OLS
-----------------------------------------------------------
                                Est.   S.E.   t val.      p
---------------------------- ------- ------ -------- ------
(Intercept)                    -1.61   0.14   -11.32   0.00
log(Sampling_time_ESC)          0.33   0.04     8.12   0.00
log(ESC_dose)                   0.17   0.03     6.57   0.00
SexM                            0.10   0.02     4.21   0.00
Age.z                          -0.12   0.01    -8.59   0.00
I(Age.z^2)                     -0.09   0.01    -8.13   0.00
TotalMeds                      -0.17   0.02    -8.72   0.00
-----------------------------------------------------------
```

#### Confounders by association with predictors of interest

Three predictors of interest determine the question of escitalopram metabolism, measured by the escitalopram D/C ratio: the CYP2C19 genetic phenotype, the co-medication (here roughly represented by the number of medications beside escitalopram) and their interaction. We therefore look for variables associated with these predictors, as they represent possible confounders we should adjust for.

The first predictor is co-medication, i.e. the number of medications beside escitalopram. In this preliminary analysis we excluded individual with 4 co-medications as there were very few of them (no individual had more than 4 co-medications).

```
MODEL INFO:
Observations: 2870
Dependent Variable: TotalMeds/3
Type: Generalized linear model
  Family: quasibinomial
  Link function: logit

MODEL FIT:
<U+03C7>²(6) = 73.81, p = 0.00
Pseudo-R² (Cragg-Uhler) = 0.06
Pseudo-R² (McFadden) = 0.05
AIC =  NA, BIC =  NA

Standard errors: MLE
-----------------------------------------------------------
                               Est.   S.E.   t val.      p
---------------------------- ------- ------ -------- ------
(Intercept)                   -5.49   0.52   -10.57   0.00
Age                            0.09   0.01     8.14   0.00
I(Age^2)                      -0.00   0.00    -6.22   0.00
SexM                          -0.04   0.08    -0.46   0.64
log(Sampling_time_ESC)         0.17   0.13     1.28   0.20
log(ESC_dose)                  0.22   0.08     2.73   0.01
CYP2C19_phenotype             -0.10   0.04    -2.42   0.02
-----------------------------------------------------------

Estimated dispersion parameter = 0.37
```

In the co-medication model, we see (as one might have assumed) a strong association between age and co-medication, but also one between co-medication and log dose. We also see again the negative association between CYP2C19 phenotype and co-medication.

The second predictor is the CYP2C19 genetic phenotype. Here, associations were observed with both escitalopram dose and number of medications:

```
MODEL INFO:
Observations: 2872
Dependent Variable: CYP2C19_phenotype/4
Type: Generalized linear model
  Family: quasibinomial
  Link function: logit

MODEL FIT:
<U+03C7>²(5) = 5.74, p = 0.00
```

```
Pseudo-R² (Cragg-Uhler) = 0.00
Pseudo-R² (McFadden) = 0.00
AIC =  NA, BIC =  NA

Standard errors: MLE
--------------------------------------------------------
                               Est.   S.E.   t val.      p
---------------------------- ------- ------ -------- ------
(Intercept)                   -0.51   0.21    -2.43   0.02
Age                            0.00   0.00     0.56   0.57
SexM                           0.04   0.04     1.28   0.20
log(Sampling_time_ESC)         0.02   0.06     0.26   0.79
log(ESC_dose)                  0.17   0.04     4.49   0.00
TotalMeds                     -0.07   0.03    -2.62   0.01
--------------------------------------------------------

Estimated dispersion parameter = 0.2
```

(we use a logistic binomial model here to account for the bounds of the outcome variable). As one may expect if the dose is adapted to the genotype, the dose increased with more active phenotypes. However, more active phenotypes also received less co-medication, a finding whose meaning is not clear. Individuals with high metabolic activity might receive more co-medications to compensate loss of efficacy, not less. We did not consider this association in the analysis that follows.

The possible causal relationships are summarized in the DAG below.

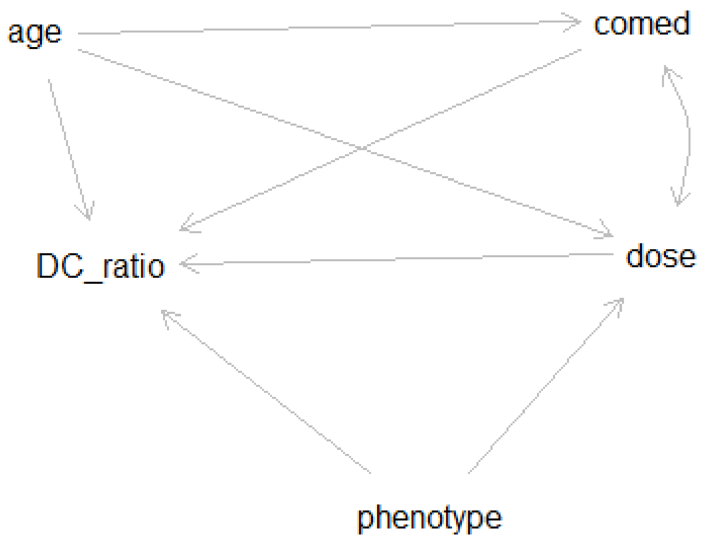


This DAG tells us that the relationship between phenotype and D/C ratio is conditionally independent (estimable without bias) if one adjusts for age, co-medication, and dose, and that the relationship between co-medication and D/C ratio is conditionally independent when adjusting for age, dose, and phenotype. In conclusion, it is essential to adjust for age and dose to model the relationship between phenotype and/or co-medication and D/C ratio (adjustment set from dagitty: age, dose). Note that this analysis is valid for direct effects (we do not want effects of dose to be included in the effect of phenotype on D/C ratio). However, this assumes that no lurking confounds that appreciably influence both dose and D/C ratio are present (conditioning on a mediator). For this reason, sensitivity analyses without dose as covariate are indicated. Furthermore, lurking confounders affecting both co-medication and D/C ratio make the effect of co-medication unidentifiable.

We did not include the relationship between phenotype and co-medication in the DAG, as we considered it a false positive. However, its inclusion does not change the adjustment set (not shown here for brevity) and the need for caution in the use of dose as a covariate.

### Exploration of data

In this preliminary exploration, we used a simple linear model with D/C ratio as outcome variable (dose to concentration), and log dose, log sampling time, sex, age (linear and quadratic) and the total number of medications as predictors.

#### Distribution of residuals

One issue in this dataset is the presence of very high D/C ratio values, which would imply extreme induction in some individuals. One possible explanation of these very high D/C ratio values is that patients were taking medications intermittently or at lower doses. Because the D/C ratio estimate takes into account dose, if the patients took less medication than assumed, it will look as if they have a high metabolic capacity. One approach to assess this possibility is to look at both mother substance and metabolite plasma concentrations. Individuals with high metabolic capacity should have high metabolite concentrations and low mother substance concentrations. Individuals who take little medication, in contrast, should have low metabolite concentrations. In the plot below, we plot the log concentrations of the mother substance and the metabolite, coloring the observations by the magnitude of the residuals.

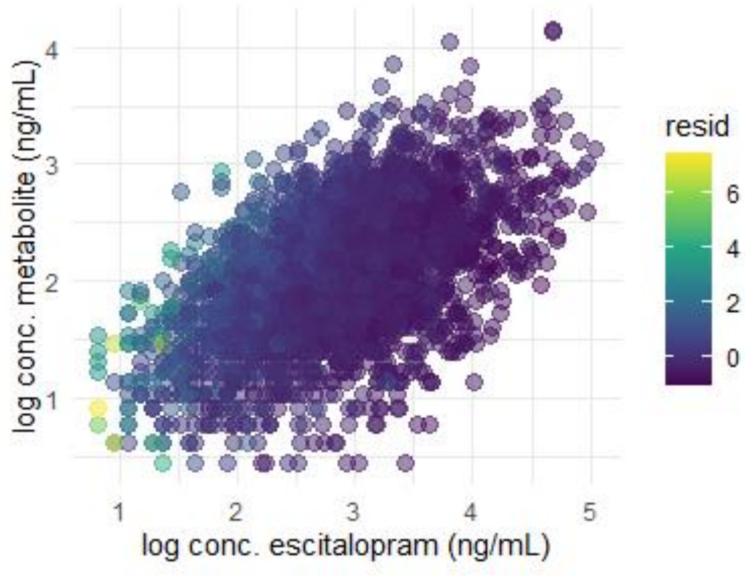

We can see that the distribution of the residual magnitude in this plot is systematic. Many individuals with high residuals (green-yellow) have low concentration of both escitalopram and its metabolite, as expected by discrepancies between the dose they were assumed to take and the one that was effectively taken (recall that our D/C ratio measure is the dose to concentration ratio, and as such makes no use of the measurement of the metabolite concentration).

Below, we plot the residual magnitude on the log product of the concentrations (of the mother substance and the metabolite), which summarizes positions along the main diagonal line in the cloud of the previous plot. In this plot, we see these high residual D/C ratio cases departing from the linear fit selectively at the low values of the predictor (that is, when both the concentration of the mother substance and the metabolite were low, as we would expect by failures in taking the medication at prescribed doses). In the plot, color codes the prescribed dose.

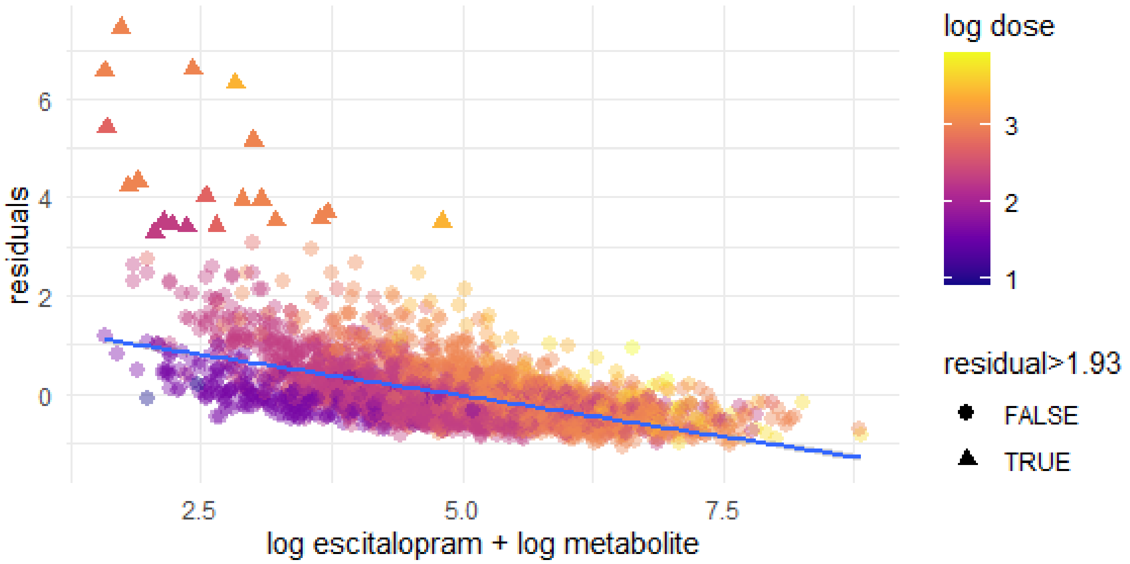


We used this model to identify observations to exclude from the analysis. The residual standard error of this model was 0.48. We therefore excluded observations with a residual larger than 4 times the residual standard error of this model, i.e. larger than 1.93 from the data (leading to the exclusion of 20 individuals). These observations were plotted as triangles in the plot above. Below, the plot of the observed D/C ratio vs. fitted D/C ratio is much improved. Before the removal, the range of observed values included values over 8, while now it goes only just over 4.

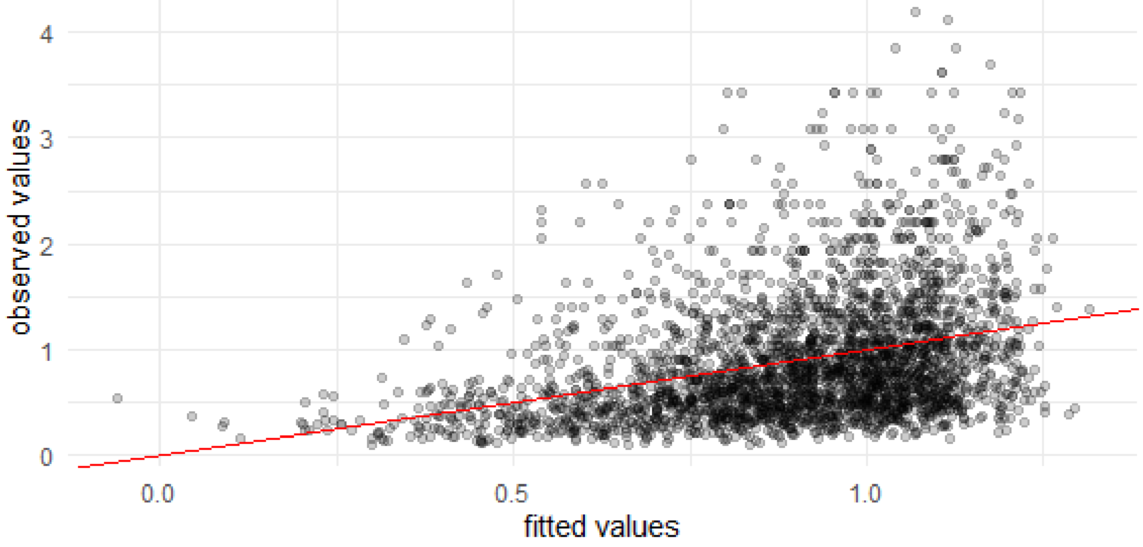


Note, however, that the variance still increases with the fitted D/C ratio values. For this reason, we will consider log-transformed models in the analyses.

### Descriptive analysis (after removal of N=21 observations)

#### Demographics and escitalopram dose

Age, sex, and dose (for 2852 observations):

| Variable | mean | median | std. dev. | range |
|---|---|---|---|---|
| age | 45.7545582 | 42 | 19.7913546 | 14-101 |
| female sex | 0.6437588 | (N = 1836) | | |
| dose | 14.0015663 | 10 | 6.2738332 | 2.5-50 |

#### Genotypes and genetic phenotypes

Genotypes were distributed with the following frequencies (in percent):

| alleles | frequency % |
|---|---|
| *1/1* | 38.91 |
| *1/17* | 24.23 |
| *1/2* | 22.76 |
| *1/3* | 0.17 |
| *1/4* | 0.10 |
| *17/17* | 4.11 |
| *2/17* | 6.23 |
| *2/2* | 2.47 |
| *2/3* | 0.03 |

| alleles | frequency % |
|---|---|
| *2/4* | 0.10 |
| *4/*17 | 0.14 |

Below is the assignment of genotype to phenotype groups and the number of cases in each combination. The columns index the phenotype groups, from 0 to 4 in increasing order of metabolic capacity. The rows are the allele combinations.

```
           0    1    2    3    4
 *1/*1      0    0 1118    0    0
 *1/*17     0    0    0  696    0
 *1/*2      0  654    0    0    0
 *1/*3      0    5    0    0    0
 *1/*4      0    3    0    0    0
 *17/*17    0    0    0    0  118
 *2/*17     0  179    0    0    0
 *2/*2     71    0    0    0    0
 *2/*3      1    0    0    0    0
 *2/*4      3    0    0    0    0
 *4/*17     0    4    0    0    0
```

#### Medication and co-medication

After the exclusion of N=21 individuals, there were 817 individuals with detected co-medication, and 2035 without. The columns in the table below are the number of co-medications.

```
   0    1    2    3    4
2035  656  146   13    2
```

In percent,

```
    0     1     2     3     4
71.35 23.00  5.12  0.46  0.07
```

We now look at the individual co-medications again in an incidence matrix.

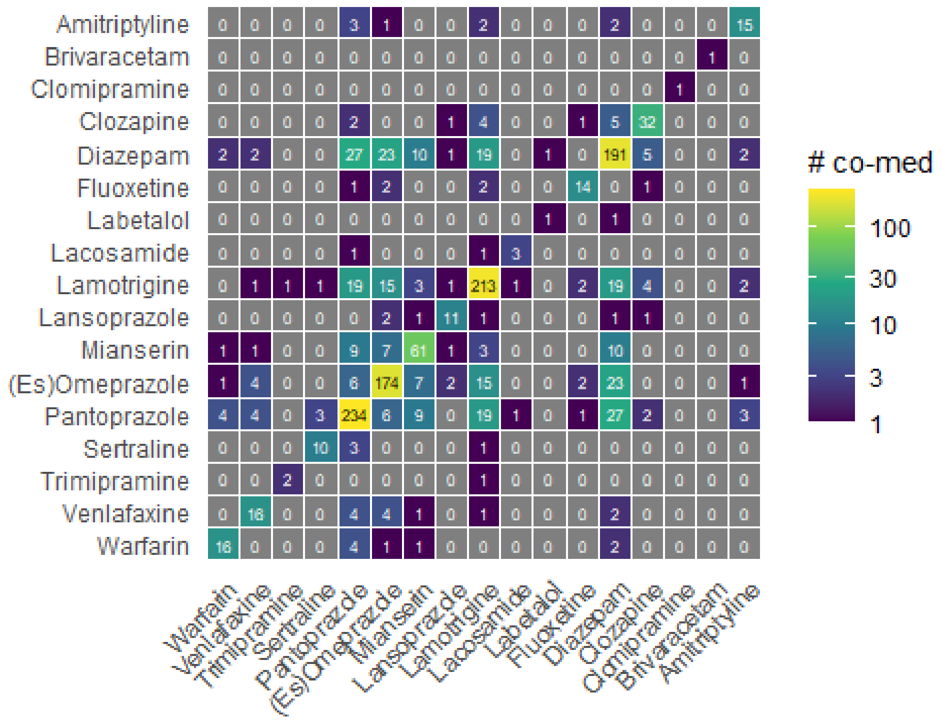


The partition of co-medication by phenotype was the following (rows, phenotype from 0 to 4, columns, number of meds):

| phenotype | count | no meds | 1 med | 2 meds | 3 meds | 4 meds |
|---|---|---|---|---|---|---|
| PM | 75 (2.6%) | 54 (72%) | 19 (25.3%) | 2 (2.7%) | NA | NA |
| IM | 845 (29.6%) | 573 (67.8%) | 210 (24.9%) | 55 (6.5%) | 5 (0.6%) | 2 (0.2%) |
| NM | 1118 (39.2%) | 807 (72.2%) | 257 (23%) | 50 (4.5%) | 4 (0.4%) | NA |
| RM | 696 (24.4%) | 514 (73.9%) | 145 (20.8%) | 34 (4.9%) | 3 (0.4%) | NA |
| UM | 118 (4.1%) | 87 (73.7%) | 25 (21.2%) | 5 (4.2%) | 1 (0.8%) | NA |

In the study, the hypothesis that will be tested is that the fractional activity scores of the medication are the predictor of competitive inhibition. For each individual in the sample, the fractional activity scores were computed from the literature and summed for the co-medications received by the individual (co-medication load). The distribution of the co-medication loads for participants receiving co-medication is shown here.

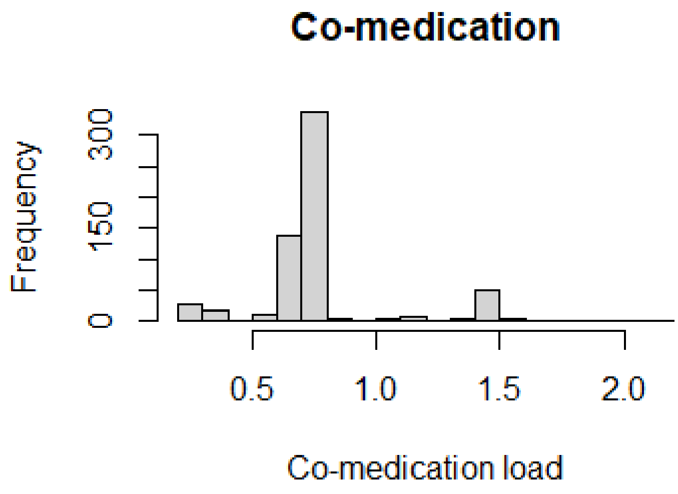


From the histogram, it is apparent that there are very sparse data to estimate effects of co-medication load over 1.5.

The histogram of the D/C ratio in the sample shows a strongly skewed distribution, as already shown in the previous plots.

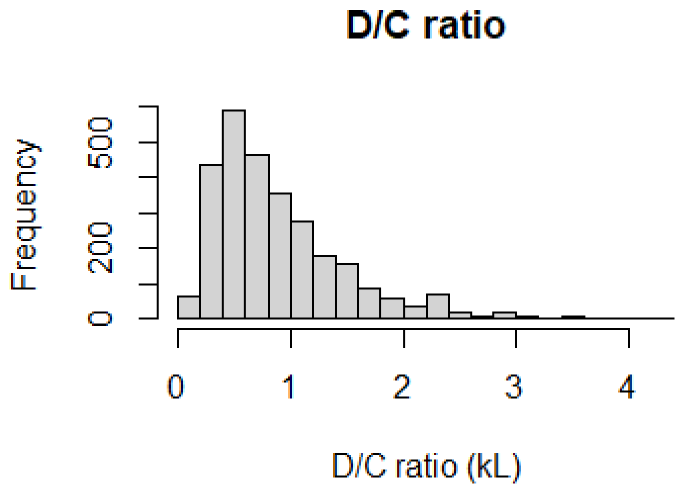


### Model parametrization

#### Data centering

Because the model will include many interactions, we will re-center the variables that enter the interactions so that the main effects remain interpretable.

#### CYP2C19 genetic phenotype

For the genetic phenotype, we want to leave the score of the PM at zero, so that the variables that enter in an interaction with the phenotype will have coefficients that estimate their effect in the PM group. These estimates (i.e., the coefficients of the main effects) will concern effects on metabolism other than those mediated by CYP2C19, as there is no CYP2C19 in PMs.

#### Age

We will standardize age, so that effects refer to the average age. This is 45.75, with a range from 14 to 101.

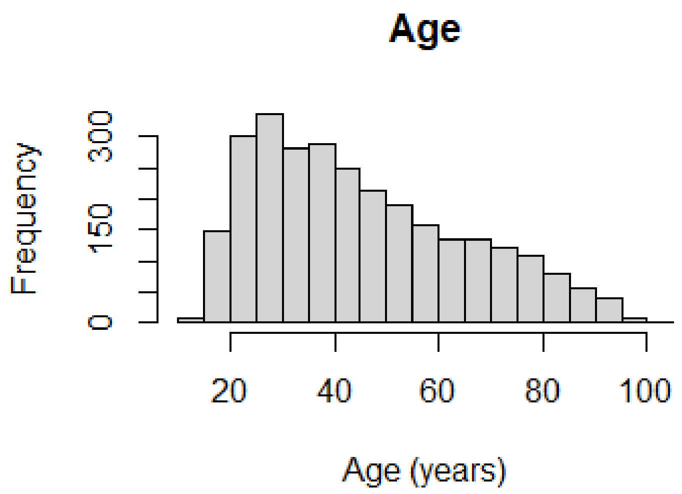


Looking at its distribution, we find all ages represented across the number of medications (an important predictor of interest), although the severely polymedicated are older:

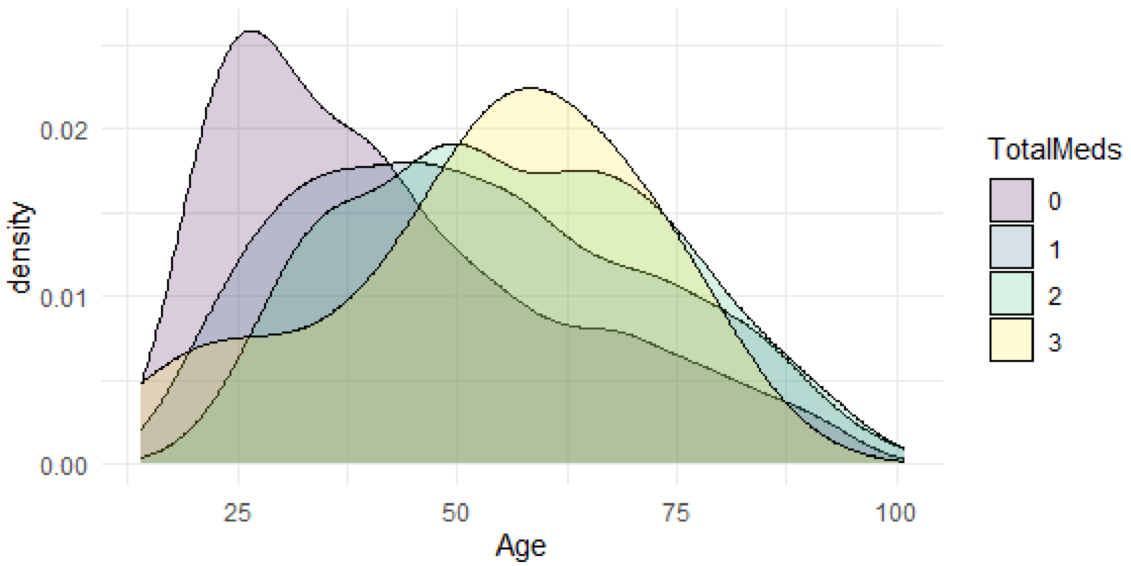


#### Escitalopram dose

We find that the most common escitalopram dose was 10 mg (as one may expect), and the most common sampling time was 24.0. We therefore prepared predictors centered at these values.

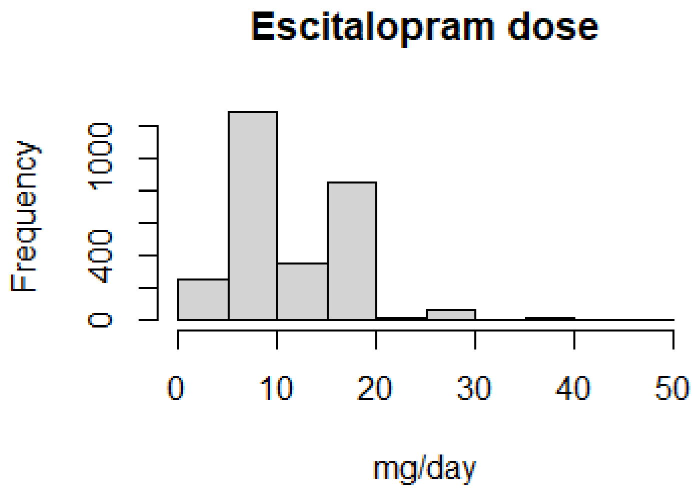


```
2.5     5  7.5     8    10    11 12.5    15 17.5    20    25    30    35    40    50
  2   257   10     1 1267     1     1   356     2   846    16    66     1    22     4
```

#### Activity scores

After identifying the confounders for the model, we take a look at the effect of the genetic phenotypes on D/C ratios. We also excluded the individuals that, according to the previous analysis, may have not taken medication as prescribed. Our working hypothesis is that it will be possible to model the D/C ratios as linear in the activity scores of CYP2C19. The idea is to establish the activity scores in the non co-medicated and to see how these scores may be used to predict D/C ratios in the co-medicated with a linear relationship.

The activity score of PM will be zero, given that there is no active enzyme in PMs. The activity score of NM will be unity, so that coefficients of the activity score give the effect on D/C ratio of moving from PM to NM. We gave an activity score of 0.5 to IM, given that it possesses one of two possible inactivating alleles.

There remains to determine the activity scores of RM and UM, which are not as well characterized. We will set them in the non-medicated such that the relationship with D/C ratio is linear. Recall that a question of interest is the effect of co-medication on this relationship. Setting a linear baseline in the non-medicated allows us to examine the emergence of non-linearities in the medicated.

When considering the data for the non co-medicated, the activity score of extensive metabolizers does not look very much different from the normal metabolizers. We therefore set the activity score for RM and UM to 1.15 and 1.3. With these activity scores, the D/C ratio increased approximately linearly in individuals without co-medication (panel A on the left; the plot below uses a loess scatterplot smoother to allow a possible non-linear fit, but one can see that the smoother produces a straight line nevertheless). The box-plots at the RM and UM phenotype groups are slightly below the fit, due to the fact that median and quantiles used in box-plots discount large outlying observations.

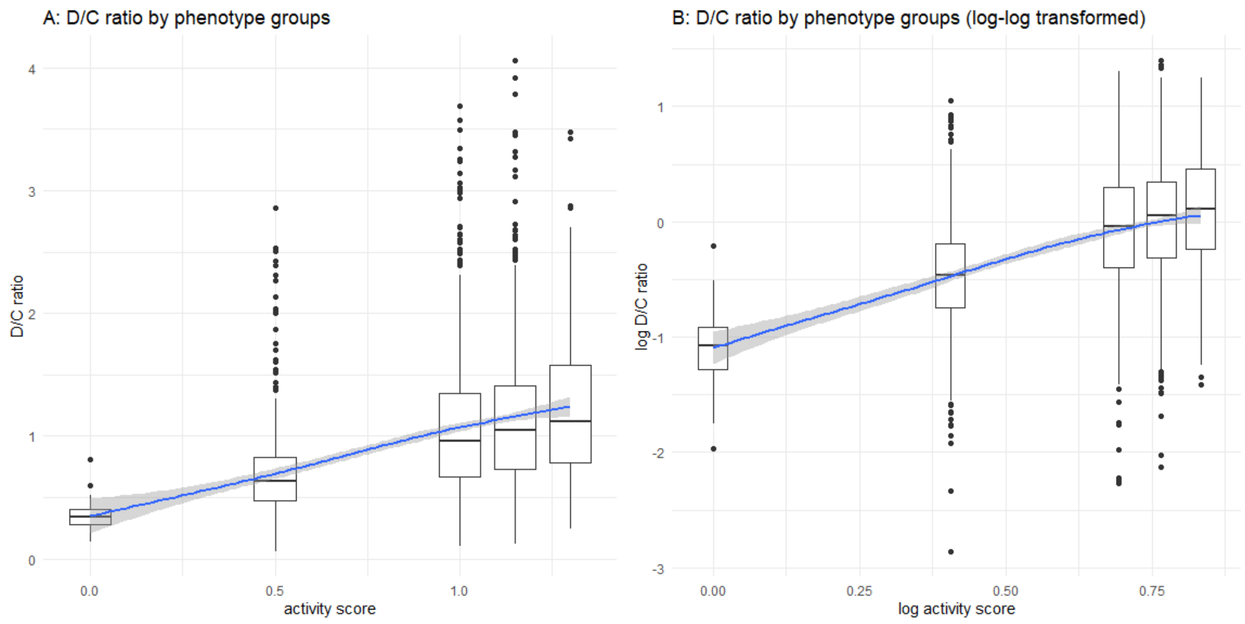


Panel B on the right shows the log-log transformed D/C ratio estimates. While not perfectly linear, the deviations from linearity are small. We used log activity scores, because the activity scores (non transformed) gave a much stronger curvature.

We may want to verify that the activity scores are still appropriate for log-log-transformed D/C ratio estimates. In the plot below, we combine the fit for the untransformed D/C ratio with one from the log-transformed D/C ratio on the log-transformed activity scores (in red).

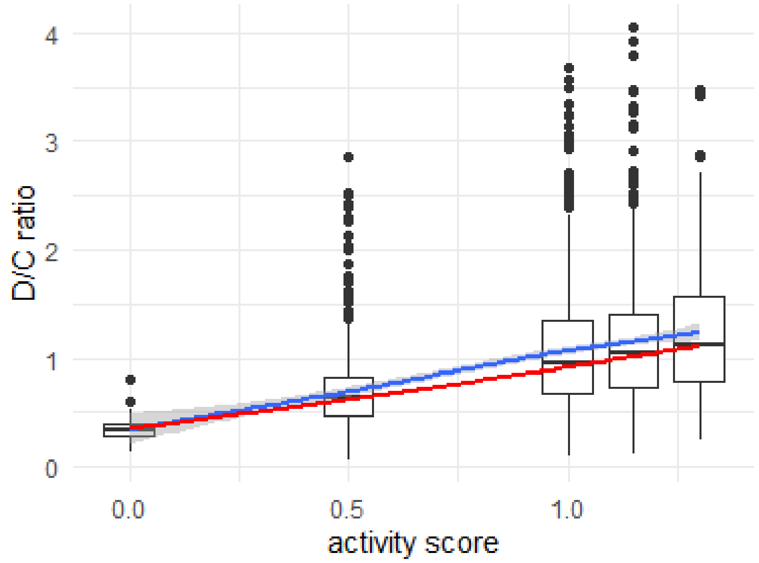


The comparison shows that the two approaches do not differ much in the extent to which they fit the data. The log-log transformed fit gives more credence to smaller observation, giving a fit that is closer to the median of the observations than the linear fit (a known property of models on log-transformed outcomes).

The $R^2$ on the original scale was 0.1433 for the linear model and 0.1430 for the log model after smearing correction, confirming that the two specifications perform similarly in this subset.

In summary, the same set of covariates is appropriate for both the linear and log-log transformed model, and the log activity scores work well in the log-transformed data.

The required preprocessing of the data is implemented in Preproc.R.
