## Supplemental file 2 for "Pharmacogenetic phenoconversion modeling of drug–drug–gene interactions on CYP2C19 activity: effects of comedication by genotype on escitalopram concentrations"

### Supplementary results 2: Models of co-medication load

Roberto Viviani, Institute of Psychology, University of Innsbruck; Psychiatry and Psychotherapy III, University of Ulm

2025-04-12

In this markdown, we analyze D/C ratios of escitalopram in genotyped individuals and with selected co-medication. Our working assumption is that the D/C ratio is a proxy of clearance values good enough to be used in lieu of them in the linear phenoconversion model. To quantify inhibition from co-medication, we will use estimates of the fractional contribution of CYP2C19 to the metabolism of co-medication from the literature (Drücker & Brockmöller 2020). In each data point, inhibition was modelled as the sum of these estimates of fractional contribution for the co-medications in that observation (*co-medication load*). This choice models competitive inhibition: the larger the role of CYP2C19 in the metabolism of a co-medication, the stronger the potential inhibition of CYP2C19.

Our first working hypothesis was that like clearance ratios (Stingl et al. 2022), D/C ratios can be modelled linearly in the activity scores and that linearity is preserved under inhibition. As shown in Figure 1 of the main text, this means that the effect of inhibition may be modelled as the interaction between activity scores and co-medication, and that the extent of the interaction represents a proportional decrease in D/C ratios (a multiplicative effect). The interaction flattens the slope of the activity score relationship while pivoting on the PM, where there is no CYP2C19 to inhibit. Hence, the lines fanning from the PM pivot encode a proportional rescaling of the activity score slope in the non-co-medicated. For example, in Figure 1, a negative interaction coefficient equal to half the coefficient of the activity scores means a reduction of 50% of non-co-medicated D/C ratios in all phenotype groups relative to those in PMs. This is a weaker claim than assuming that the effect of activity scores on D/C ratios is the same as on clearance, but establishes that both types of changes are proportional.

After verifying linearity, our second working hypothesis will be that the proportional changes due to co-medication may be linearly related to proportional changes in clearance ratios produced by the co-medication, as indexed by the fractional contribution of CYP2C19 to the metabolism of the co-medication. This is a weaker assumption than a definite functional relationship between D/C and clearance ratios. As in Cox regression, where relative hazards are estimated without specifying the baseline hazard, the quantity of interest is a proportional change that may be estimated as invariant to the form that baseline ratios take. Even when the functional relationship between activity scores and clearance or D/C ratios in the non-co-medicated is not identical, proportional changes in the two arising from the same cause may be linearly related. We will set out to verify this empirically in the Supplementary file 3. Here, we will show that the fractional contributions from the literature may be used to predict inhibition in CYP2C19 in the D/C ratios.

#### Visualization of data

When visualizing the D/C ratio by phenotype group, it is apparent that the dispersion of the data differed across these groups (figure, panel A). This is consistent with data showing increasing variance at higher values, given that the phenotype groups are predictive of enzyme activity. We also see the usual skewness of positive-only data of this kind.

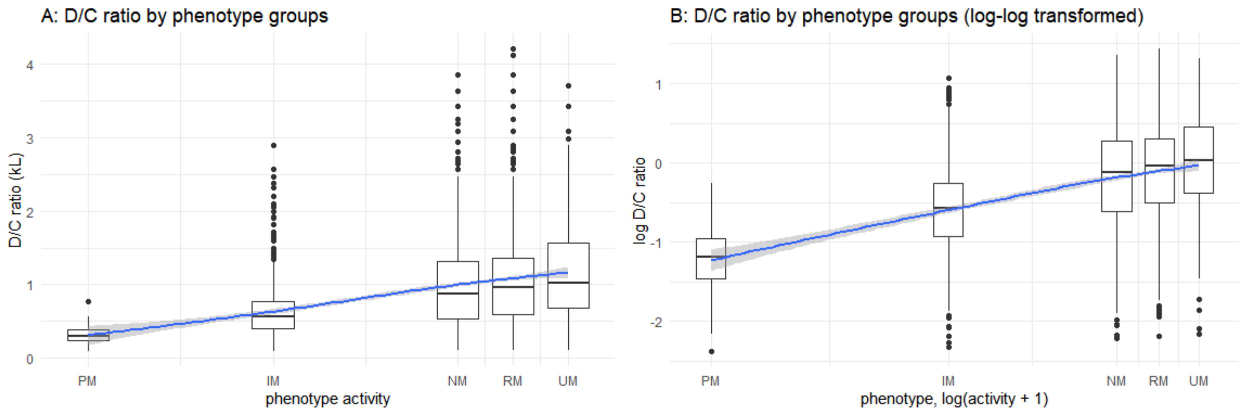


A main issue in this section is how to model this variability when using co-medication load and phenotypes as predictors. To stabilize variance, a natural approach would be to put the model in log space, either by log-transforming the data (panel B of the figure) or by using a generalized linear model (GLM) with the appropriate link function. Here, however, we need to consider the possible mechanisms of this increasing variance more closely. One mechanisms is that factors affecting metabolism by acting on the available protein will have effects amplified by its amount. This gives a multiplicative effect, as in a model in log space, even in a linear model. Given that the amount of available protein is determined by the genetic phenotype, the increasing variance can also be modelled by predictors interacting with the genetic phenotype. This is indeed what the linear phenoconversion model foresees for gene-drug interactions (Viviani, Berres, & Stingl 2024). Since there is no protein in the PM, the variance there is lowest; and as the phenotype activity increases, so does the possible variability of the metabolic capacity (see Figure 1 in the main text).

More generally, because the genotype determines the amount of functional protein, the possibility of protein function being modified dynamically is larger in high expression genetic phenotypes, giving rise to as many opportunities to affect D/C ratio through an interaction with the phenotype.

In the following, we will fit a set of models to clarify the implications of these two different modelling choices (log-transformed or linear with interactions). First, we will fit a linear model with heteroscedastic residuals, modelling residual variance as a function of the activity scores to verify its relation to expected CYP2C19 activity (linear or quadratic). We will then model residual variance in a GLM respecting the power relation suggested by the first model, which will turn out to suggest modelling in log space. Finally, we will explore a gamma GLM in linear space (i.e., with an identity link) and explore its advantages.

The main result will be that the predicted metabolic capacity (D/C ratio) and the magnitude of inhibition is largely the same in all models. However, the models differ in the identification of the mechanisms through which inhibition ensues. While only one model is reported in the main text, the comparison between models reported here is key to clarifying what conclusions are robust to modelling choices and why the gamma GLM with an identity link will be used later in the Bayesian analysis in the Supplementary results 3.

#### Linear model with heteroscedastic residuals

This model implements the linear phenoconversion model directly as an interaction, as just described. A linear regression included co-medication load and its interaction with the activity scores of the genetic phenotypes as the main predictors of interest. The model also included the covariates identified in the Data exploration section:

- sex
- age, linear and quadratic (and their interaction with activity scores)
- log dose (and its interaction with activity scores)
- log sampling time (and its interaction with activity scores).

Putting together all these predictors and the constant term in a regression matrix $X$, the coefficients in $\beta$, and denoting the outcome variable D/C ratio with $y$:

$$y = X\beta + \epsilon$$

The variance of the errors $\epsilon$ was modelled as increasing with the activity score of the phenotype groups according to a power law, with the value of the exponent being estimated from the data:

$$\text{Var}(\epsilon) = \sigma^2\left(\delta_1 + AS^{\delta_2}\right)^2$$

(cf. Pinheiro and Bates 2000, pp. 212-213), where *AS* is the activity score of the genetic phenotype. In this formula, $\sigma\delta_1$ gives the estimate of the standard deviation of the PMs (if one sets *AS* to zero, $Var(\epsilon) = \sigma^2\delta_1^2$). The variance then increases proportionally to $AS^{\delta_2}$, where $\delta_2$ is the parameter to be estimated.

To assess the adequacy of the model for the residuals, we compared it with one where the variance of the residuals was constant (OLS), and one in which the variance was estimated separately for each genetic phenotype, thus making no assumption as to the form that the variance takes.

```
        Model df      AIC      BIC    logLik   Test  L.Ratio p-value
fit_LS0     1 14 4476.417 4559.734 -2224.208                        
fit_LS1     2 16 4030.756 4125.975 -1999.378 1 vs 2 449.6612  <.0001
fit_LS2     3 18 4031.777 4138.898 -1997.888 2 vs 3   2.9789  0.2255
```

As expected, the model with a power model of heteroscedastic variance fitted the data very much better than the model with uniform variance (*p* < 0.001), as was apparent from the plots of the data. The third variance model that fits separate variances in each genetic phenotype group was not significantly better. We conclude that the second model was a reasonable fit to the data.

#### Model results

The main hypothesis was that the interaction between the co-medication load and activity scores would be negative as an effect of the inhibition of CYP2C19 on metabolism increasing proportionally to the original DME activity. A second question of interest was the effect of increasing co-medication in the PMs. As hypothesized in Stingl & Viviani (2025), co-medication may decrease the metabolism of the substrates in the PMs by inhibiting other enzymes.

```
Generalized least squares fit by REML
  Model: DC_ratio ~ CYP2C19Act * (TotalFC + Age.z + Age.z.2 + Smplt_ESC.log.c +      ESC_dose.
log.c) + Sex.c
  Data: data_selection
       AIC      BIC    logLik
  4030.756 4125.975 -1999.378

Variance function:
 Structure: Constant plus power of variance covariate
 Formula: ~CYP2C19Act
 Parameter estimates:
    const     power
0.2521753 0.9999982

Coefficients:
                                Value  Std.Error   t-value p-value
(Intercept)                 0.3388406 0.01902893 17.806606  0.0000
CYP2C19Act                  0.7908060 0.02970835 26.618983  0.0000
TotalFC                    -0.0516210 0.03870445 -1.333722  0.1824
Age.z                      -0.0329076 0.01572696 -2.092431  0.0365
Age.z.2                    -0.0043623 0.01164745 -0.374524  0.7080
Smplt_ESC.log.c            -0.0252730 0.04022581 -0.628279  0.5299
ESC_dose.log.c             -0.0217495 0.02509901 -0.866546  0.3863
Sex.c                      -0.0641034 0.01556265 -4.119056  0.0000
CYP2C19Act:TotalFC         -0.2390622 0.05792206 -4.127308  0.0000
CYP2C19Act:Age.z           -0.0536078 0.02279157 -2.352087  0.0187
CYP2C19Act:Age.z.2         -0.0612592 0.01784554 -3.432746  0.0006
CYP2C19Act:Smplt_ESC.log.c  0.3837220 0.06206176  6.182906  0.0000
CYP2C19Act:ESC_dose.log.c   0.1445726 0.03899319  3.707639  0.0002

 Correlation:
                           (Intr) CYP2C19Ac TotlFC Age.z  Ag.z.2 S_ESC. ESC_..
CYP2C19Act                 -0.755
TotalFC                    -0.371  0.278
Age.z                       0.316 -0.234    -0.312
Age.z.2                    -0.584  0.450     0.056 -0.579
Smplt_ESC.log.c             0.286 -0.212     0.013 -0.031 -0.002
ESC_dose.log.c             -0.384  0.302     0.198  0.024  0.134  0.017
Sex.c                      -0.180  0.069     0.038  0.033 -0.019 -0.056  0.115
CYP2C19Act:TotalFC          0.295 -0.334    -0.820  0.255 -0.044 -0.014 -0.166
CYP2C19Act:Age.z           -0.254  0.295     0.258 -0.813  0.469  0.026 -0.021
CYP2C19Act:Age.z.2          0.464 -0.612    -0.042  0.442 -0.782  0.002 -0.115
CYP2C19Act:Smplt_ESC.log.c -0.222  0.287    -0.013  0.026 -0.001 -0.766 -0.005
CYP2C19Act:ESC_dose.log.c   0.306 -0.413    -0.147 -0.027 -0.106 -0.009 -0.757
                           Sex.c  CYP2C19A:T CYP2C19Ac:A. CYP2C19A:A..
CYP2C19Act
```

```
TotalFC
Age.z
Age.z.2
Smplt_ESC.log.c
ESC_dose.log.c
Sex.c
CYP2C19Act:TotalFC         -0.035
CYP2C19Act:Age.z           -0.015 -0.285
CYP2C19Act:Age.z.2         -0.021  0.046     -0.528
CYP2C19Act:Smplt_ESC.log.c  0.061  0.007     -0.061        0.009
CYP2C19Act:ESC_dose.log.c  -0.070  0.112      0.050        0.158
                           CYP2C19A:S
CYP2C19Act
TotalFC
Age.z
Age.z.2
Smplt_ESC.log.c
ESC_dose.log.c
Sex.c
CYP2C19Act:TotalFC
CYP2C19Act:Age.z
CYP2C19Act:Age.z.2
CYP2C19Act:Smplt_ESC.log.c
CYP2C19Act:ESC_dose.log.c   0.007

Standardized residuals:
       Min         Q1        Med         Q3        Max
-1.9237809 -0.6568216 -0.1897632  0.4059536  6.3646321

Residual standard error: 0.4503538
Degrees of freedom: 2852 total; 2839 residual
```

In this model,

- (Intercept) is the estimated D/C ratio of escitalopram (in kL) at 24h (at trough) in PM individuals of average age taking 10mg/day of escitalopram and no co-medication;
- *CYP2C19Act* is the effect on D/C ratios of CYP2C19 activity scores (the difference in D/C ratios between NMs and PMs) in individuals of average age in the without co-medication group receiving a dose of 10mg/day of escitalopram, with plasma levels assessed at 24h;
- *TotalFC* is the effect of co-medication load in PM individuals;
- *Age.z* and *Age.z.2* model the effects of increasing age in PM individuals (linear and quadratic terms);
- *ESC_dose.log.c* models the effect of log dose on the D/C ratio in PM individuals;
- *Smplt_ESC.log.c* is the effect of log time at which plasma was taken, also in PM individuals;
- *Sex.c* is the effect of sex;
- *CYP2C19Act:TotalFC* is the interaction of co-medication load and CYP2C19 activity scores, i.e., the change in the effect of activity scores per one unit of co-medication load;
- *CYP2C19Act:Age.z* and *CYP2C19Act:Age.z.2* model the interaction between age and CYP2C19 activity scores;
- *CYP2C19Act:ESC_dose.log.c* models the interaction between log dose and the activity scores;
- *CYP2C19Act:Smplt_ESC.log.c* is the interaction between the log sampling time and activity scores.

Furthermore, the ‘power’ estimate in the ‘Variance function’ section is the exponent $\delta_2$ of the model of residuals, reported earlier.

In this model, estimates of the D/C ratio of escitalopram at 24h (at trough) in NMs taking 10mg/day may be obtained by summing the coefficients of the intercept (D/C ratio in PMs) and of activity scores (the difference in D/C ratios between NMs and PMs). This gives a volume of 1.12965 kL.

This model tells us the following. First, as hypothesized, the interaction between inhibition and activity scores was negative and significant ($p < 0.001$). In the cases without co-medication, the coefficient of the activity score was 0.79. This coefficient decreased by 0.24 for a hypothetical co-medication that is fully metabolized by CYP2C19, i.e. by about 30%.

Second, in PMs (activity score 0), the effect of co-medication load failed to reach stringent significance thresholds ($p = 0.09$, one tailed), but was negative. While co-medication load for CYP2C19 is not the best predictor to assess inhibition of other enzymes, it strongly correlates with the number of co-medications taken. For this reason, the negative coefficient suggests that there may be an effect in PMs whose mechanism differs from the effect identified by the interaction between co-medication load and activity scores.

In contrast, age was a significant predictor of D/C ratios in PMs ($p = 0.036$). This may be interpreted as being due to a general effect on metabolic capacity other than on CYP2C19. Age was also significant in the interaction with the activity scores ($p = 0.019$) that is, like co-medication it acted on the relationship between genetic phenotype and D/C ratios by flattening the activity score line. Age also showed a strong quadratic component in the interaction, meaning that the effect of age increased with increasing age (test of combined linear and quadratic effects via likelihood ratio, $\chi_4^2 = 133.53$, $p < 0.001$).

Below, we show linear fits of the phenotype group on D/C ratios separately for co-medication load groups (the lines are slightly shifted to the right for increasing co-medication to align them with the box-plots). The co-medication load was split into four equally spaced groups to show the effect on the slope of the phenotype groups. One can see that the slope of these linear fits flattens with increasing co-medication load. However, there were just 3 observations in the highest load group, which makes it not indicative.

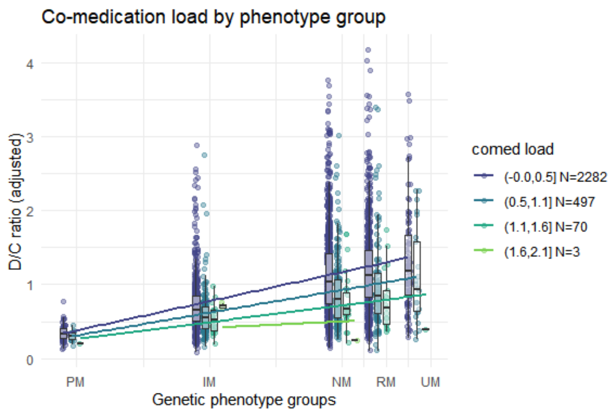


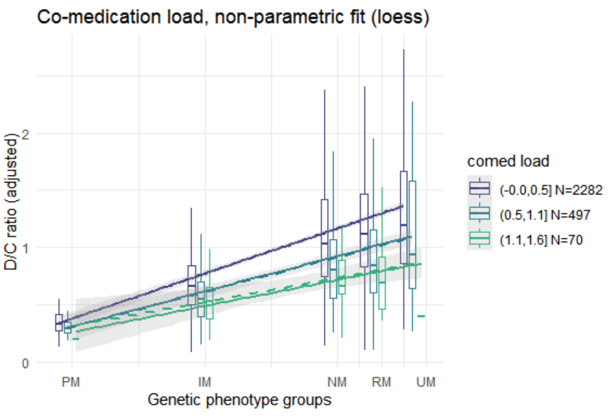

A main tenet of the linear phenoconversion model is that the proportionality of the effects of co-medication inhibition maintains the linearity of the relationship between activity scores and clearance (here replaced by D/C ratios). We shall test this formally by adding a quadratic term to the model. In the right part of the figure above, we used a scatterplot smoother on the data of the groups with increasing co-medication load. In these latter groups, if the relationship between activity scores and inhibition deviates from linearity, it will be shown in the smoother as a curve. One can see that the scatterplot smoother is almost identical to the fitted line. In the third group, there are few observations in the PM and UM groups, such that the confidence intervals of the fitted curve overlap with the fitted line. The fourth group is too small to be fitted with a scatterplot smoother.

In the following, we plotted the interaction between genetic phenotype and age. The plot shows the strong quadratic effects of age (left panel). In this more balanced groups, the linearity of the fit with the scatterplot smoother is apparent (right panel).

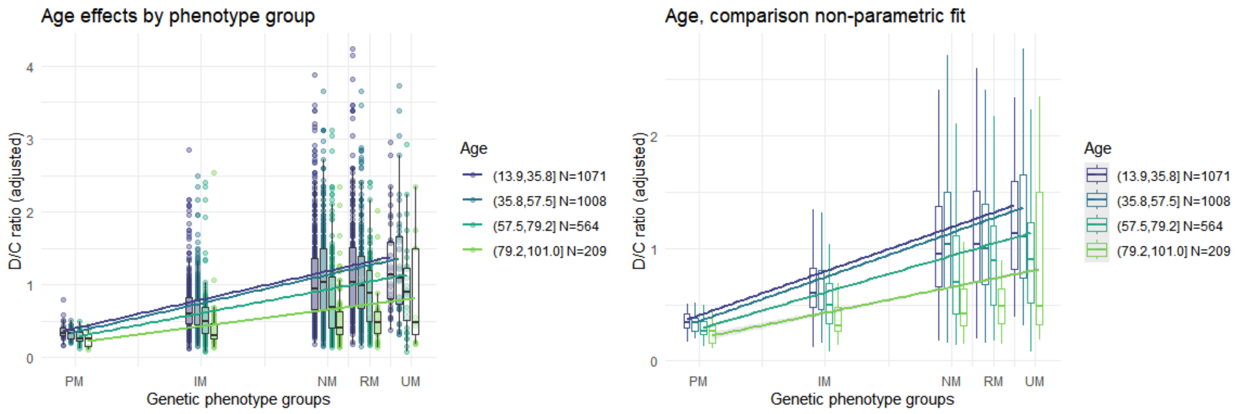


There was a very large amount of unexplained variation in the D/C ratio in the phenotype groups except PMs, meaning that our capacity to predict metabolic capacity was moderate at best.

#### Linearity of the phenoconversion

The previous plot showed that the scatterplot smoother fit had confidence intervals overlapping with the linear fit. To quantify this linearity more precisely, we fit a quadratic term of the activity scores. The quadratic term would be able to detect stronger effects of co-medication in IMs relative to a linear prediction (as in the downward shift model of phenoconversion, giving a positive quadratic coefficient) as well as stronger inhibition in UMs (non-linearity of the relationship between activity scores and D/C ratios, giving a negative coefficient).

The models showed that no significant deviations from linearity were present in the data (quadratic term of activity scores in the interaction with co-medication load: $t$ = -0.6635, $p$ = 0.51).

Another issue is the possible non-linearity of the interaction with the co-medication load, as its effect may saturate at high loads. We do not see any sign of this, however. This might be due to the moderate levels of co-medication in the data.

However, also in this case there was no significant result (quadratic term of co-medication load in the interaction with activity scores: $t = -0.0376$, $p = 0.97$).

#### Sensitivity analysis

The previous models were refitted without log dose and its interaction with the activity scores as covariates. The conclusions were the same.

Specifically, the interaction between activity scores and co-medication load remained significant ($t = -4.228$, $p < 0.001$). The quadratic term of the activity scores in the interaction with co-medication load remained non-significant ($t = -0.4118$, $p = 0.68$).

#### Model diagnostics

The estimated $\delta_2$ parameter in this model was unity (0.9999982), implying that the variance increased quadratically with the increasing activity scores.

The plot of Pearson residuals vs. the fitted values shows a fairly uniform amplitude distribution, except for observations at very low predicted D/C ratio. However, the residuals are still skewed, with much longer tails on the upper end, as the heteroscedastic model cannot redress this. This means that only a model that takes into account the skewness of residuals can deliver appropriate confidence intervals.

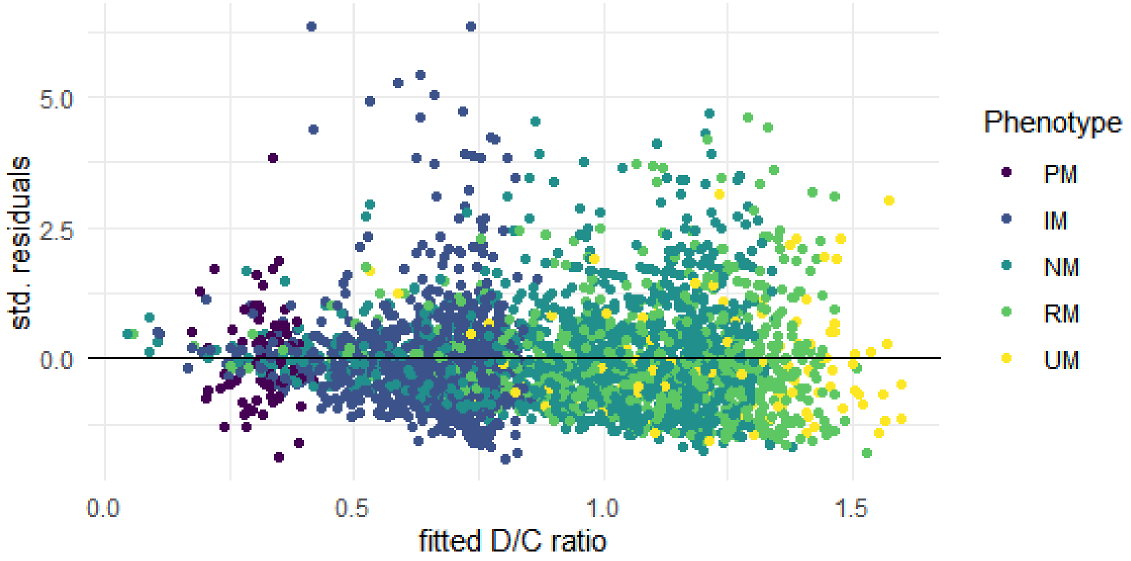


#### Linear regression: conclusion

In conclusion, we see that first, most predictors, except for sex, tended to affect the D/C ratio by interacting with the activity score, i.e. their effect was amplified with the available functional protein. Furthermore, this effect maintained the linearity of the relationship between activity scores and D/C ratio, as in the linear phenoconversion model.

Second, it appears that there are predictors that we do not know about but may affect the D/C ratio through the same mechanism. Even if we included almost all our predictors in the interaction with the activity scores, the variance was still increasing with the value of these scores. There were many observations that departed from the predicted D/C ratio, and most of them were located in the cone obtained by changing the slope of the activity score predictor, i.e the error of the prediction broadly increased with the activity score. These affect both directions, as there were individuals in the UM group that have D/C ratio levels of PMs.

Third, the residuals were still very skewed. This means that confidence intervals from this model cannot reflect the variability of the underlying data.

One final observation is that the variance in the PM was low in absolute terms. This means that factors that affect measured D/C ratio beyond those allowed by the presence of the functional protein, such as measurement error, seem to have small effects on the data. An alternative possibility is that the errors in measurement are multiplicative: at higher D/C ratios, the measurement becomes less precise. This alternative possibility is difficult to disentangle with statistical means from that of dynamic changes in D/C ratios.

From a purely data-driven modelling perspective, the skewness of the residuals remains and their variance scaling with the square of the activity scores (the main predictor of main activity) strongly suggests the use of a log-transform in the model. This would be the most common approach to modelling this kind of data. We turn to the log-transformed model in the next section.

#### Gamma generalized linear model, log link

We will fit the same structural model as in the previous section, but we now resort to a gamma generalized linear model (GLM) with a log link and log-transformed activity scores (the log of the scores after adding a unity), following the observations on the power of residual variance in the preliminary analysis (for a discussion of this kind of models in pharmacokinetic data, see Salway & Wakefield 2008). Writing predictors and coefficients in matrix form as $X$ and $\beta$ as before, and denoting D/C ratio by $y$, our model becomes

$$\log(y) \sim \text{gamma}(X\beta, \phi)$$

where $\phi$ is the shape parameter of the gamma distribution.

```
Call:
glm(formula = DC_ratio ~ CYP2C19Act.log * (TotalFC + Age.z +
    I(Age.z^2) + Smplt_ESC.log.c + ESC_dose.log.c) + Sex.c, family = Gamma(link = "log"),
    data = data_selection)

Deviance Residuals:
    Min       1Q   Median       3Q      Max
```

```
-1.6608  -0.4354  -0.1193   0.2110   2.6509

Coefficients:
                                Estimate Std. Error t value Pr(>|t|)
(Intercept)                    -0.990526   0.061677 -16.060  < 2e-16 ***
CYP2C19Act.log                  1.627833   0.097396  16.714  < 2e-16 ***
TotalFC                        -0.144269   0.110022  -1.311 0.189869
Age.z                          -0.081962   0.046484  -1.763 0.077969 .
I(Age.z^2)                     -0.053145   0.036427  -1.459 0.144686
Smplt_ESC.log.c                -0.009833   0.131467  -0.075 0.940384
ESC_dose.log.c                  0.022468   0.081880   0.274 0.783797
Sex.c                          -0.081007   0.022129  -3.661 0.000256 ***
CYP2C19Act.log:TotalFC         -0.316306   0.175437  -1.803 0.071501 .
CYP2C19Act.log:Age.z           -0.022503   0.072470  -0.311 0.756190
CYP2C19Act.log:I(Age.z^2)      -0.058357   0.057687  -1.012 0.311806
CYP2C19Act.log:Smplt_ESC.log.c  0.534542   0.206173   2.593 0.009572 **
CYP2C19Act.log:ESC_dose.log.c   0.148588   0.128470   1.157 0.247537
---
Signif. codes:  0 '***' 0.001 '**' 0.01 '*' 0.05 '.' 0.1 ' ' 1

(Dispersion parameter for Gamma family taken to be 0.3162367)

    Null deviance: 1159.91  on 2851  degrees of freedom
Residual deviance:  789.98  on 2839  degrees of freedom
AIC: 2833.5

Number of Fisher Scoring iterations: 5
```

We see here that many interactions of the linear model are no longer significant (including the interaction between activity scores and co-medication load). This is consistent with the dependency of residual variance on the activity scores. Because the activity score is a strong predictor of the available protein and this model expressed multiplicative effects, the percentage change in D/C ratio postulated by the linear phenoconversion model may be expressed directly by the activity scores, instead of an interaction with them.

This model may therefore be simplified by removing all interactions except the one with sampling time,

```
Call:
glm(formula = DC_ratio ~ CYP2C19Act.log * Smplt_ESC.log.c + TotalFC +
    Age.z + Age.z.2 + ESC_dose.log.c + Sex.c, family = Gamma(link = "log"),
    data = data_selection)

Deviance Residuals:
    Min       1Q   Median       3Q      Max
-1.6575  -0.4363  -0.1206   0.2119   2.7902

Coefficients:
                               Estimate Std. Error t value Pr(>|t|)
(Intercept)                    -0.93380    0.04376 -21.337  < 2e-16 ***
CYP2C19Act.log                  1.53456    0.06527  23.510  < 2e-16 ***
Smplt_ESC.log.c                 0.01689    0.13083   0.129 0.897279
TotalFC                        -0.33169    0.03123 -10.621  < 2e-16 ***
Age.z                          -0.09590    0.01260  -7.613 3.63e-14 ***
Age.z.2                        -0.08722    0.01048  -8.326  < 2e-16 ***
ESC_dose.log.c                  0.11485    0.02406   4.773 1.91e-06 ***
Sex.c                          -0.08110    0.02211  -3.668 0.000249 ***
```

```
CYP2C19Act.log:Smplt_ESC.log.c  0.49402    0.20507   2.409 0.016058 *
---
Signif. codes:  0 '***' 0.001 '**' 0.01 '*' 0.05 '.' 0.1 ' ' 1

(Dispersion parameter for Gamma family taken to be 0.3163016)

    Null deviance: 1159.9  on 2851  degrees of freedom
Residual deviance:  792.8  on 2843  degrees of freedom
AIC: 2836.2

Number of Fisher Scoring iterations: 5
```

This model is more parsimonious, and provides massive evidence for inhibition from medication ($t > 10$ in absolute terms). According to this model, one unit of fractional contribution decreases D/C ratios by about 28% relative to unmedicated levels. This value is very similar to the one obtained with the linear regression model. Also estimates of the D/C ratio at trough are similar (1.13868 kL).

The plots of the fits for co-medication load and age groups confirm them to be similar to those of the linear model with heteroscedastic residuals (the scatterplot smoother uses a generalized linear model with a log link and splines as predictors, as in the model itself).

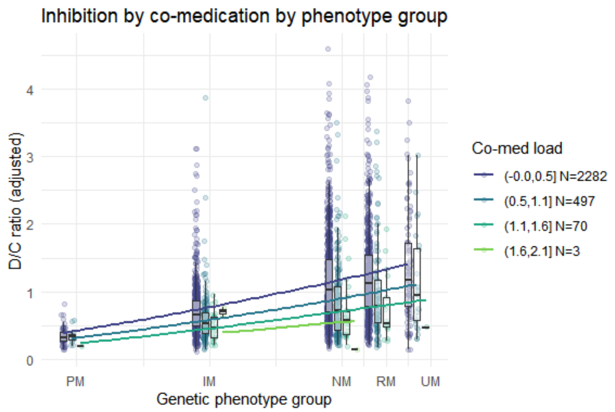


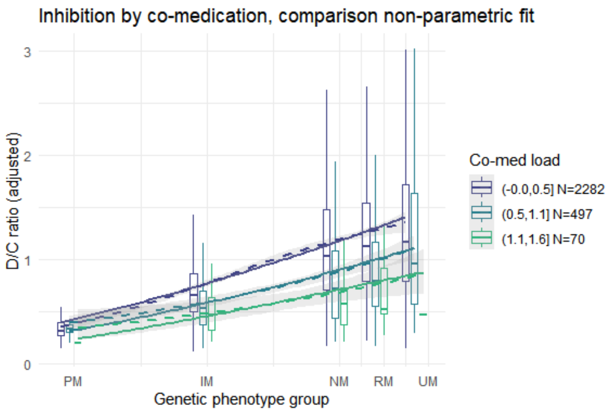


There is a slight curvature for the individuals with no co-medication load (this was investigated in the Exploratory section of the analysis, showing it to deviate little from a linear fit). The curvature becomes even less marked the higher the co-medication load. Even without interaction, the fit shows progressive flattening because of the multiplicative effects induced by the log link.

#### Linearity of the phenoconversion

In this context, the question may be reframed as asking if the relationship changes with increasing inhibition, so as to make it independent of if the relationship was linear to start with. Also in this model, we see no evidence that the form of the relationship between D/C ratio and activity scores changes with increasing inhibition.

```
Call:
glm(formula = DC_ratio ~ CYP2C19Act.log * (TotalFC + Smplt_ESC.log.c) +
    I(CYP2C19Act.log^2) * TotalFC + Age.z + Age.z.2 + ESC_dose.log.c +
```

```
    Sex.c, family = Gamma(link = "log"), data = data_selection)

Deviance Residuals:
    Min       1Q   Median       3Q      Max
-1.6480  -0.4366  -0.1189   0.2100   2.6991

Coefficients:
                               Estimate Std. Error t value Pr(>|t|)
(Intercept)                    -1.07810    0.07308 -14.753  < 2e-16 ***
CYP2C19Act.log                  2.10374    0.28266   7.443 1.30e-13 ***
TotalFC                        -0.01381    0.19694  -0.070 0.944115
Smplt_ESC.log.c                 0.01146    0.13079   0.088 0.930191
I(CYP2C19Act.log^2)            -0.49889    0.27333  -1.825 0.068070 .
Age.z                          -0.09535    0.01259  -7.572 4.96e-14 ***
Age.z.2                        -0.08817    0.01047  -8.425  < 2e-16 ***
ESC_dose.log.c                  0.11570    0.02406   4.809 1.60e-06 ***
Sex.c                          -0.08082    0.02208  -3.660 0.000257 ***
CYP2C19Act.log:TotalFC         -0.83328    0.77989  -1.068 0.285403
CYP2C19Act.log:Smplt_ESC.log.c  0.50351    0.20497   2.457 0.014088 *
TotalFC:I(CYP2C19Act.log^2)     0.45636    0.74182   0.615 0.538482
---
Signif. codes:  0 '***' 0.001 '**' 0.01 '*' 0.05 '.' 0.1 ' ' 1

(Dispersion parameter for Gamma family taken to be 0.3154242)

    Null deviance: 1159.91  on 2851  degrees of freedom
Residual deviance:  790.23  on 2840  degrees of freedom
AIC: 2832.5

Number of Fisher Scoring iterations: 5
```

#### Model diagnostics

We now examine the residuals with deviance plots.

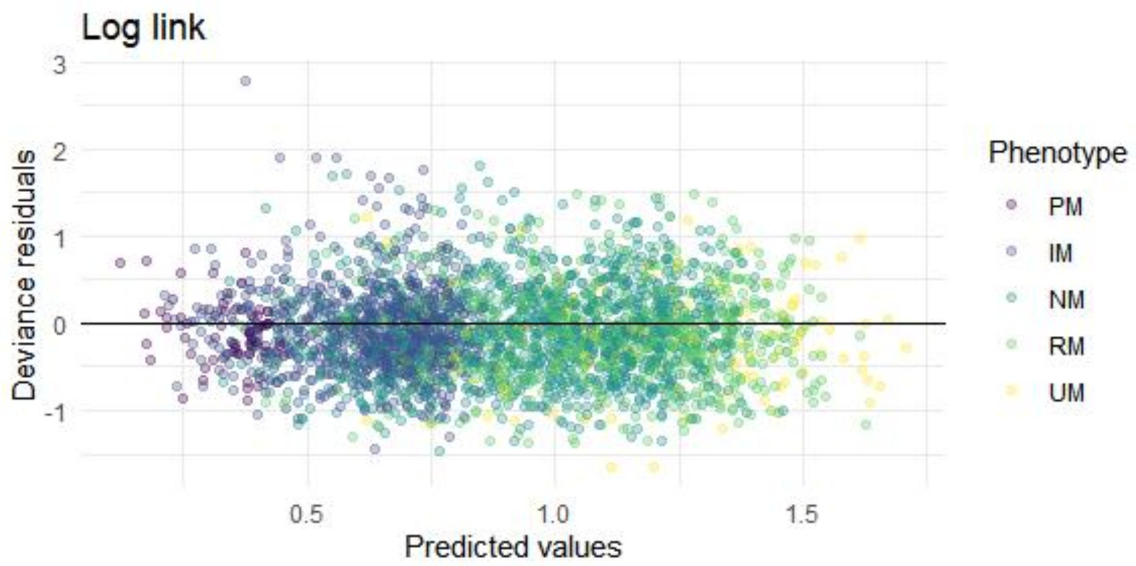


The distribution of deviance residuals is much more uniform than in the linear model, and shows no skewness, suggesting that the log-transformed model has addressed the issue of the distribution of residuals well. The outlying observation turns out to be the IM with a

TotalFC of 0.76 with large D/C ratio values (visible in the plots below but not looking too implausible).

##### Issues with this model

While the model shows a good fit, it should be noted that it does not identify the increasing variance of the residuals as an interaction with the activity scores. This interaction is a plausible mechanism through which factors affect D/C ratios by acting on the enzyme, at least for co-medication. In a model with interaction, it is possible to distinguish between effects in the PM (due to potential effects of co-medication on enzymes other than CYP2C19) and effects on the CYP2C19. The model forces every predictor to act proportionally on D/C ratios, and since D/C ratios are strongly predicted by the phenotype, it models this interaction implicitly. Note also that coefficients in the log space are not as immediately interpretable as coefficients of a linear model.

#### Gamma generalized linear model, identity link

To maintain the identifiability of main effects and interactions of activity scores, we fitted a gamma GLM with an identity link (see McCullagh and Nelder, pp. 294-295). Originally meant to model sum of squares in variance component models, this is a combination rarely seen in applied work. Here, the rationale was that the model should adapt to the skewed shape of the residuals while maintaining the linear predictor, thus allowing us to differentiate between effects of co-medication at the PM and their interaction with activity scores:

$$y \sim \text{gamma}(X\beta, \phi)$$

where the predictors in $X$ are again those of the linear model with heteroscedastic residuals.

```
Call:
glm(formula = DC_ratio ~ CYP2C19Act * (TotalFC + Age.z + I(Age.z^2) +
    Smplt_ESC.log.c + ESC_dose.log.c) + Sex.c, family = Gamma(link = "identity"),
    data = data_selection)

Deviance Residuals:
    Min       1Q   Median       3Q      Max
-1.6579  -0.4409  -0.1188   0.2156   2.4863

Coefficients:
                            Estimate Std. Error t value Pr(>|t|)
(Intercept)                 0.324807   0.024244  13.397  < 2e-16 ***
CYP2C19Act                  0.773475   0.032811  23.574  < 2e-16 ***
TotalFC                    -0.015680   0.039006  -0.402 0.687720
Age.z                      -0.033081   0.021018  -1.574 0.115615
I(Age.z^2)                 -0.003731   0.013122  -0.284 0.776149
Smplt_ESC.log.c            -0.037226   0.053260  -0.699 0.484635
ESC_dose.log.c              0.004412   0.031560   0.140 0.888832
Sex.c                      -0.060426   0.016791  -3.599 0.000325 ***
CYP2C19Act:TotalFC         -0.268323   0.049824  -5.385 7.82e-08 ***
CYP2C19Act:Age.z           -0.048486   0.027017  -1.795 0.072811 .
```

```
CYP2C19Act:I(Age.z^2)      -0.045984   0.016851  -2.729 0.006395 **
CYP2C19Act:Smplt_ESC.log.c  0.320584   0.067989   4.715 2.53e-06 ***
CYP2C19Act:ESC_dose.log.c   0.112274   0.042278   2.656 0.007961 **
---
Signif. codes:  0 '***' 0.001 '**' 0.01 '*' 0.05 '.' 0.1 ' ' 1

(Dispersion parameter for Gamma family taken to be 0.3128927)

    Null deviance: 1159.91  on 2851  degrees of freedom
Residual deviance:  794.66  on 2839  degrees of freedom
AIC: 2851.1

Number of Fisher Scoring iterations: 10
```

The comparison favours the log link model (lower AIC and deviance). However, we now recover the inference on the interactions. The interaction between co-medication load and activity scores was negative and significant ($p < 0.001$). In the cases without co-medication, the coefficient of the activity score was 0.77. This coefficient decreased by 0.27 for a hypothetical co-medication that is fully metabolized by CYP2C19, i.e a decrease by about 35%, similar to the linear regression model. Also similar to the linear regression model are the estimates of D/C ratios at trough (1.09828 kL).

In PMs, age failed to reach significance ($p = 0.116$). Age was significant especially in the quadratic term of the interaction with activity scores, as decreases in D/C ratios were particularly marked in older individuals ($p = 0.006$).

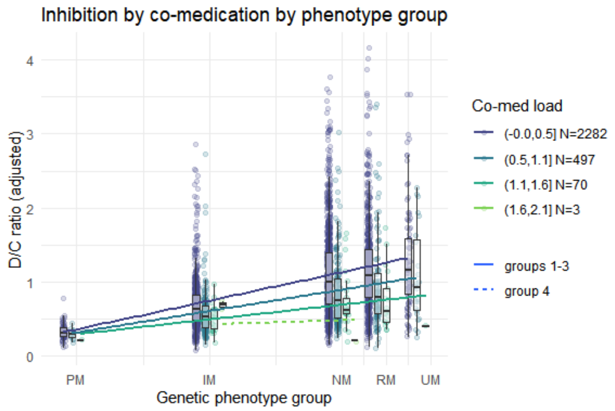


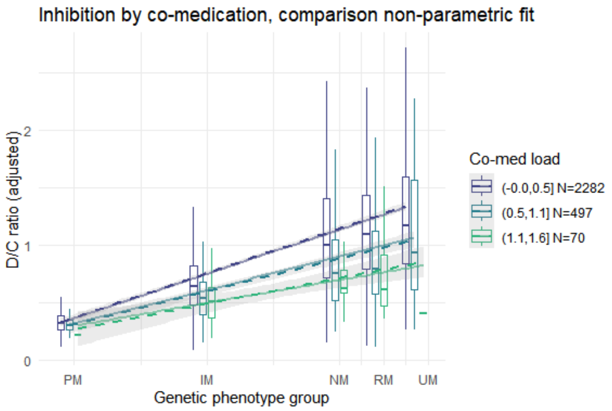


We can see that in the fitted data, the slopes of the activity scores flattened with increasing co-medications, as in the linear phenoconversion model. As the value and range of variation in D/C ratios gets smaller in the high co-medication load, the slope was also flatter.

The effect of age also shows a flattening of the curve with an acceleration after 60.

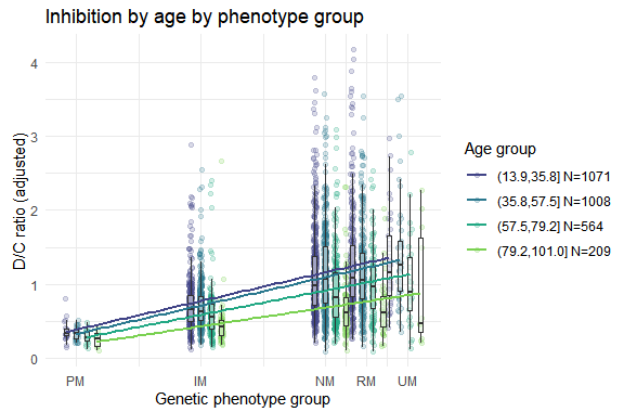


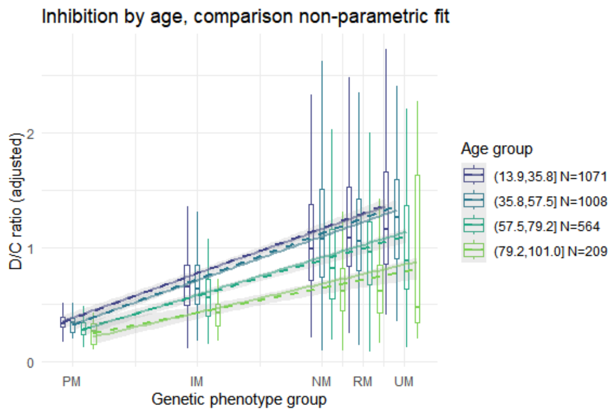


```
Warning: glm.fit: algorithm did not converge

Analysis of Deviance Table

Model 1: DC_ratio ~ CYP2C19Act * (TotalFC + Smplt_ESC.log.c + ESC_dose.log.c) +
    Age.z + I(Age.z^2) + Sex.c
Model 2: DC_ratio ~ CYP2C19Act * (TotalFC + Age.z + I(Age.z^2) + Smplt_ESC.log.c +
    ESC_dose.log.c) + Sex.c
  Resid. Df Resid. Dev Df Deviance      F    Pr(>F)
1      2841     808.42
2      2839     794.66  2   13.763 21.994 3.323e-10 ***
---
Signif. codes:  0 '***' 0.001 '**' 0.01 '*' 0.05 '.' 0.1 ' ' 1

Analysis of Deviance Table

Model 1: DC_ratio ~ CYP2C19Act * (TotalFC + Smplt_ESC.log.c + ESC_dose.log.c) +
    CYP2C19Act:(Age.z + I(Age.z^2)) + Sex.c
Model 2: DC_ratio ~ CYP2C19Act * (TotalFC + Age.z + I(Age.z^2) + Smplt_ESC.log.c +
    ESC_dose.log.c) + Sex.c
  Resid. Df Resid. Dev Df Deviance      F  Pr(>F)
1      2841     796.57
2      2839     794.66  2   1.9138 3.0583 0.04712 *
---
Signif. codes:  0 '***' 0.001 '**' 0.01 '*' 0.05 '.' 0.1 ' ' 1
```

Age was a significant phenoconverting agent (linear and quadratic effects, $F = 21.994$, $p < 0.001$), that is, like co-medication it acted on the relationship between genetic phenotype and D/C ratios by flattening the activity score line. However, it was also a significant inhibitor in PMs ($F = 3.058$, $p = 0.047$). This means that the inhibitory effects of age cannot be modelled on the range bounded below by PM metabolic activity.

##### *Common plot meds and age*

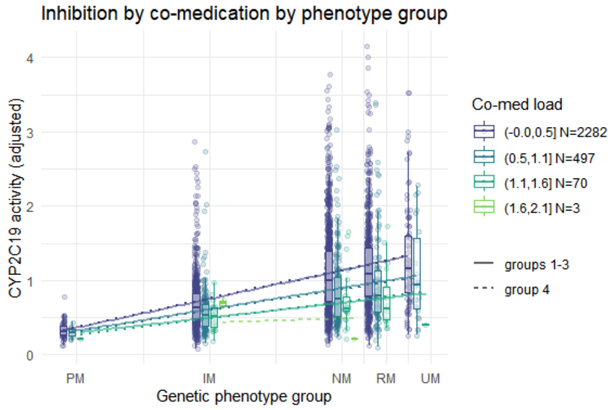


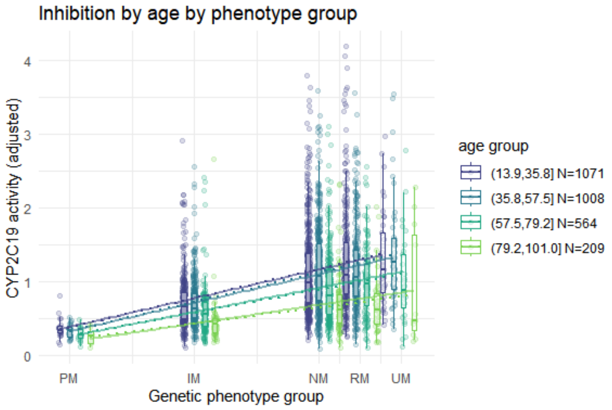


#### Linearity of phenoconversion

We repeat here the quadratic fits for this model.

The models showed that no significant deviations from linearity were present in the data (quadratic term of activity scores in the interaction with co-medication load: $t$ = -0.4184, $p$ = 0.68).

Another issue is the possible non-linearity of the interaction with the co-medication load, as its effect may saturate at high loads. We do not see any sign of this, however (quadratic term of co-medication load in the interaction with activity scores: -0.0115, $p$ = 0.92). This might be due to the moderate levels of co-medication in the data.

#### Sensitivity analysis

We fit here a model that does not include log dose as confounder, showing again a significant interaction between activity scores and co-medication load:

```
Call:
glm(formula = DC_ratio ~ CYP2C19Act * (TotalFC + Age.z + I(Age.z^2) +
    Smplt_ESC.log.c) + Sex.c, family = Gamma(link = "identity"),
    data = data_selection)

Deviance Residuals:
    Min       1Q   Median       3Q      Max
-1.6781  -0.4505  -0.1169   0.2094   2.4474

Coefficients:
                            Estimate Std. Error t value Pr(>|t|)
(Intercept)                 0.319830   0.022862  13.990  < 2e-16 ***
CYP2C19Act                  0.817669   0.030654  26.674  < 2e-16 ***
TotalFC                    -0.006651   0.039430  -0.169 0.866068
Age.z                      -0.030830   0.021113  -1.460 0.144330
I(Age.z^2)                 -0.002809   0.013065  -0.215 0.829760
Smplt_ESC.log.c            -0.052881   0.053969  -0.980 0.327244
Sex.c                      -0.065513   0.016928  -3.870 0.000111 ***
CYP2C19Act:TotalFC         -0.266162   0.050517  -5.269 1.48e-07 ***
CYP2C19Act:Age.z           -0.053699   0.027114  -1.981 0.047742 *
```

```
CYP2C19Act:I(Age.z^2)      -0.059221   0.016561  -3.576 0.000355 ***
CYP2C19Act:Smplt_ESC.log.c  0.330810   0.068955   4.797 1.69e-06 ***
---
Signif. codes:  0 '***' 0.001 '**' 0.01 '*' 0.05 '.' 0.1 ' ' 1

(Dispersion parameter for Gamma family taken to be 0.3179675)

    Null deviance: 1159.91  on 2851  degrees of freedom
Residual deviance:  802.61  on 2841  degrees of freedom
AIC: 2876.9

Number of Fisher Scoring iterations: 8
```

Also the quadratic term in this model is not significant, as before (quadratic term of activity scores in the interaction with co-medication load: $t = -0.5833$, $p = 0.56$).

These analyses confirm the conclusions of the previous model.

#### Model diagnostics

We now examine the residuals with deviance plots.

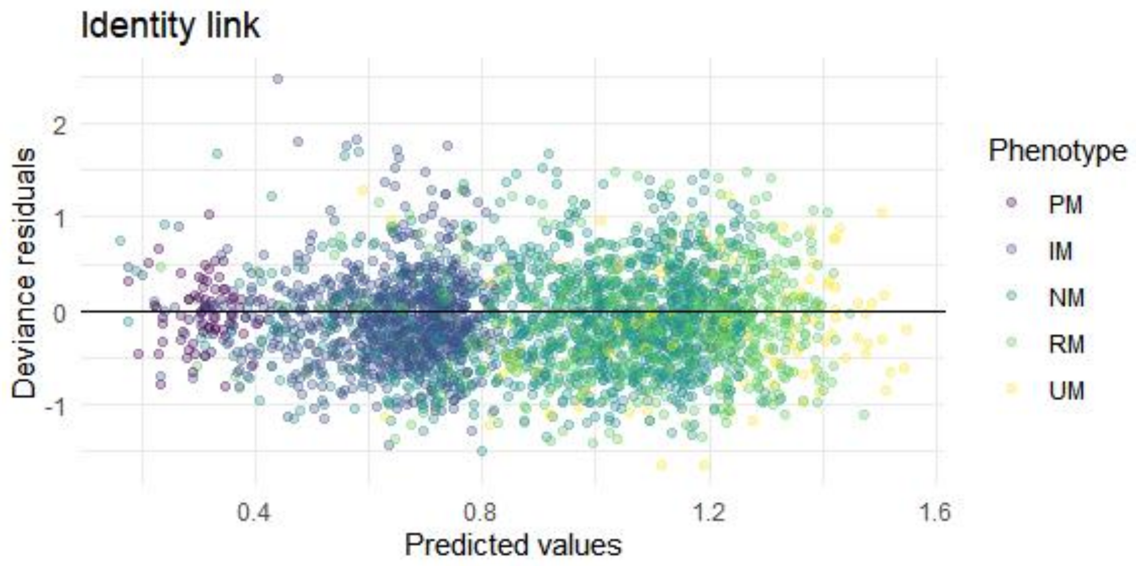


The distribution of deviance residuals is similar in both the log link and identity link models, showing that this latter was successful in modelling the shape of residuals.

### Conclusion

All models gave essentially the same fits of D/C ratios, and proportional estimates of progressive inhibition were similar. However, the models differed in two respects:

- the linear and the gamma GLM with identity link identified effects of inhibition at the PM and in CYP2C19 through the interaction with activity scores;
- the gamma GLMs with log and identity link were capable of modelling skewness in the residual variability.

While the gamma GLM with log link gives adequate and parsimonious predictions of D/C ratios, the gamma GLM with identity link is important to separate effects at the PM and in CYP2C19. This is therefore the model we chose in the main text and in the Supplementary results 3.

#### Limitations

Because the data do not contain information on individuals taking more than four medications, these models cannot be informative about the cumulative effects of polypharmacy for more than 4 co-medications. Indeed, the data become very sparse already for 2 co-medications, so that one cannot model the saturation of inhibition from these data. In the linear predictor without saturation, the slope of the activity scores will become negative at some point when more co-medications are added. This cannot happen, and is a limitation of all these models, also of the log-transformed model. This model ensures that there can be no negative predictions, but a negative activity scores slope would still produce D/C ratios estimates in UMs that are lower than in PMs, which is not biologically plausible.
