## Supplemental file 3 for "Pharmacogenetic phenoconversion modeling of drug–drug–gene interactions on CYP2C19 activity: effects of comedication by genotype on escitalopram concentrations"

### Supplementary results 3: Bayesian model of inhibition of individual co-medications

Roberto Viviani, Institute of Psychology, University of Innsbruck; Psychiatry and Psychotherapy III, University of Ulm

2025-04-27

#### Models of individual medications

We used here a Bayesian approach to model the amount of inhibition of CYP2C19 of individual co-medications. The Bayesian approach is required to have a stable model capable of handling medications with very few cases in the data (three co-medications were seen only once).

In this model, we fitted a predictor containing the number of co-medications (whose coefficient estimates the average effect of adding one medication) in a gamma GLM with identity link. We then added one predictor per medication, and modelled all these predictors with *t* distributions with 6 dfs (one group for the main effect, one group of coefficients for the interaction with the activity scores). These coefficients model the departure from the mean effect of adding a co-medication, thus identifying the individual inhibitory effect of each co-medication, and the *t* distribution shrinks estimates towards the mean medication effect for data with few observations. As in the non-Bayesian models, the main effect represents the effect on D/C ratio at 24h at the PM phenotype, and the interactions with the activity score, representing the change in the slope of the activity score with increasing inhibition (phenoconversion). The question of interest was the extent to which estimated phenoconversion by co-medication correlated with *in vivo* estimates of the fractional contribution of CYP2C19 of the same co-medication (Dücker & Brockmöller 2020).

The model included the same covariates as in the models of the co-medication load. This gave the following design matrix for the fixed effects:

- intercept
- activity scores
- number of co-medications (and its interaction with activity scores)
- sex
- age, linear and quadratic (and their interaction with activity scores)
- log dose (and its interaction with activity scores)
- log sampling time (and its interaction with activity scores).

Putting in $Z_1$ and $Z_2$ the incidence matrix of the presence of a co-medication and its interaction with the activity scores, respectively, we may formulate the model as follows

$$y \sim \text{gamma}(Xb + Z_1 q + Z_2 r, \phi)$$

and

$$q_j \sim t_6\left(0, \sigma_q^2\right), r_j \sim t_6\left(0, \sigma_r^2\right)$$

where *y* are the *N* observations of D/C ratios, *b* are the coefficient estimates, including the intercept, *q* is the random effect vector of co-medications at the PM, $q_j$ for $j = 1...M$, the number of co-medications, and *r* is the corresponding vector of the interaction estimates of the co-medications with the activity scores, and $\phi$ the shape parameter of the gamma distribution. The *t* distributions with 6 dfs of the effects of individual co-medication in the PM and on the activity score slope have standard deviations $\sigma_q$ and $\sigma_r$.

Estimates of effects of individual medications may be retrieved from the model as the sum of the mean effect of adding a co-medication and the specific coefficient modelling deviation from the mean effect for the medication in question.

In the model, we set moderately informative priors for $\sigma_q^2$ and $\sigma_r^2$ to facilitate convergence, based on the fact that the coefficients for medication at the PM and for the interaction with the activity scores cannot be larger than the expected D/C ratio at the PM and the coefficient of activity scores itself.

```
Inference for Stan model: anon_model.
4 chains, each with iter=6000; warmup=2000; thin=1;
post-warmup draws per chain=4000, total post-warmup draws=16000.

                      mean se_mean    sd      2.5%       50%     97.5% n_eff
act                  0.737   0.000 0.031     0.675     0.738     0.798 11846
sex                 -0.049   0.000 0.015    -0.079    -0.049    -0.019 22133
agexact             -0.063   0.000 0.025    -0.112    -0.063    -0.015 11293
age2xact            -0.036   0.000 0.016    -0.068    -0.036    -0.006  8830
dosexact             0.107   0.000 0.039     0.031     0.107     0.184 18496
timexact             0.296   0.000 0.061     0.179     0.296     0.417 16509
medsTotxact         -0.068   0.002 0.093    -0.248    -0.070     0.120  3846
medsxact[1]         -0.195   0.002 0.196    -0.606    -0.184     0.158 10328
medsxact[2]         -0.001   0.002 0.282    -0.577    -0.007     0.589 16485
medsxact[3]         -0.039   0.002 0.291    -0.663    -0.029     0.532 18757
medsxact[4]          0.204   0.002 0.206    -0.154     0.185     0.667 11371
medsxact[5]          0.027   0.002 0.104    -0.184     0.029     0.228  4613
medsxact[6]         -0.244   0.002 0.167    -0.601    -0.231     0.049  7605
medsxact[7]          0.002   0.003 0.310    -0.627     0.004     0.631 13861
medsxact[8]          0.040   0.002 0.245    -0.405     0.026     0.562 13667
medsxact[9]          0.165   0.002 0.108    -0.047     0.164     0.377  4889
medsxact[10]        -0.109   0.002 0.166    -0.439    -0.108     0.222  9919
medsxact[11]         0.221   0.002 0.142    -0.046     0.217     0.508  7994
medsxact[12]        -0.390   0.002 0.112    -0.619    -0.387    -0.179  4457
medsxact[13]        -0.057   0.002 0.106    -0.271    -0.055     0.148  4394
medsxact[14]        -0.041   0.002 0.212    -0.466    -0.041     0.391 16563
medsxact[15]         0.004   0.002 0.259    -0.500     0.000     0.547 16075
medsxact[16]         0.150   0.002 0.161    -0.150     0.143     0.486  8095
medsxact[17]         0.147   0.002 0.174    -0.178     0.138     0.519 10276
sigma_meds           0.080   0.000 0.029     0.035     0.076     0.149  4257
sigma_actxmeds       0.240   0.001 0.079     0.120     0.228     0.428  5814
shape                3.819   0.001 0.100     3.627     3.817     4.019 26044
mu                   0.341   0.000 0.023     0.297     0.340     0.388  9663
age                 -0.025   0.000 0.020    -0.064    -0.025     0.013  9459
age2                -0.010   0.000 0.013    -0.034    -0.010     0.015  7635
```

```
logDose              -0.001   0.000 0.028    -0.056    -0.001     0.055 15945
smplTime             -0.046   0.000 0.049    -0.144    -0.045     0.048 14094
medsTot              -0.017   0.001 0.062    -0.137    -0.016     0.105  3936
meds[1]               0.146   0.001 0.145    -0.103     0.133     0.467  9465
meds[2]               0.000   0.001 0.175    -0.353     0.001     0.356 16493
meds[3]               0.011   0.001 0.179    -0.347     0.008     0.380 17089
meds[4]              -0.058   0.001 0.139    -0.342    -0.054     0.208 11881
meds[5]              -0.020   0.001 0.071    -0.158    -0.021     0.123  5024
meds[6]               0.009   0.001 0.122    -0.206     0.000     0.265  6774
meds[7]              -0.057   0.001 0.177    -0.412    -0.056     0.292 14066
meds[8]              -0.029   0.001 0.160    -0.366    -0.025     0.273 14310
meds[9]              -0.031   0.001 0.070    -0.168    -0.031     0.109  5259
meds[10]              0.050   0.001 0.119    -0.175     0.047     0.297 12143
meds[11]             -0.048   0.001 0.093    -0.224    -0.049     0.141  8680
meds[12]              0.082   0.001 0.081    -0.075     0.082     0.242  4673
meds[13]             -0.002   0.001 0.072    -0.142    -0.003     0.141  4585
meds[14]              0.047   0.001 0.139    -0.223     0.042     0.335 15414
meds[15]              0.000   0.001 0.168    -0.340     0.001     0.336 15096
meds[16]             -0.034   0.001 0.127    -0.280    -0.036     0.225  9795
meds[17]             -0.039   0.001 0.140    -0.311    -0.041     0.247 11858
inhibition[1]        -0.263   0.002 0.194    -0.656    -0.256     0.103 11854
inhibition[2]        -0.070   0.003 0.295    -0.654    -0.074     0.554 13383
inhibition[3]        -0.107   0.003 0.303    -0.752    -0.100     0.492 14334
inhibition[4]         0.136   0.002 0.206    -0.229     0.120     0.586 12540
inhibition[5]        -0.041   0.000 0.062    -0.166    -0.040     0.078 19606
inhibition[6]        -0.312   0.001 0.149    -0.626    -0.304    -0.036 13287
inhibition[7]        -0.066   0.003 0.325    -0.728    -0.064     0.602 11208
inhibition[8]        -0.028   0.002 0.255    -0.491    -0.045     0.518 12176
inhibition[9]         0.097   0.000 0.064    -0.031     0.097     0.220 18463
inhibition[10]       -0.178   0.001 0.160    -0.481    -0.182     0.150 16612
inhibition[11]        0.153   0.001 0.124    -0.092     0.153     0.392 18852
inhibition[12]       -0.459   0.000 0.062    -0.580    -0.458    -0.340 17053
inhibition[13]       -0.125   0.000 0.056    -0.237    -0.125    -0.016 22104
inhibition[14]       -0.109   0.002 0.218    -0.537    -0.110     0.336 15613
inhibition[15]       -0.064   0.002 0.271    -0.584    -0.073     0.506 12429
inhibition[16]        0.082   0.002 0.155    -0.220     0.079     0.395 10320
inhibition[17]        0.079   0.002 0.171    -0.249     0.074     0.428 11560
inhibition_PM[1]      0.129   0.002 0.149    -0.130     0.117     0.456  8915
inhibition_PM[2]     -0.017   0.002 0.185    -0.386    -0.015     0.356 12751
inhibition_PM[3]     -0.006   0.002 0.189    -0.380    -0.008     0.381 13137
inhibition_PM[4]     -0.075   0.001 0.144    -0.363    -0.073     0.205 10733
inhibition_PM[5]     -0.037   0.000 0.046    -0.120    -0.038     0.058 20857
inhibition_PM[6]     -0.008   0.001 0.111    -0.194    -0.020     0.238 10950
inhibition_PM[7]     -0.074   0.002 0.185    -0.442    -0.072     0.294 12219
inhibition_PM[8]     -0.046   0.002 0.168    -0.393    -0.042     0.273 12248
inhibition_PM[9]     -0.048   0.000 0.044    -0.129    -0.051     0.046 16996
inhibition_PM[10]     0.034   0.001 0.121    -0.199     0.030     0.284 15036
inhibition_PM[11]    -0.064   0.001 0.084    -0.211    -0.070     0.113 14522
inhibition_PM[12]     0.065   0.000 0.052    -0.033     0.063     0.172 16429
inhibition_PM[13]    -0.019   0.000 0.044    -0.101    -0.019     0.069 22895
inhibition_PM[14]     0.030   0.001 0.147    -0.249     0.027     0.333 13027
inhibition_PM[15]    -0.017   0.002 0.178    -0.376    -0.015     0.340 11168
inhibition_PM[16]    -0.051   0.001 0.132    -0.297    -0.056     0.225  9606
inhibition_PM[17]    -0.056   0.001 0.147    -0.336    -0.061     0.252 10139
lp__              -1340.472   0.113 6.534 -1354.239 -1340.125 -1328.607  3337
                   Rhat
act               1.000
sex               1.000
agexact           1.000
age2xact          1.000
dosexact          1.000
```

```
timexact          1.000
medsTotxact       1.001
medsxact[1]       1.001
medsxact[2]       1.000
medsxact[3]       1.000
medsxact[4]       1.000
medsxact[5]       1.001
medsxact[6]       1.001
medsxact[7]       1.000
medsxact[8]       1.000
medsxact[9]       1.001
medsxact[10]      1.000
medsxact[11]      1.000
medsxact[12]      1.001
medsxact[13]      1.001
medsxact[14]      1.000
medsxact[15]      1.000
medsxact[16]      1.000
medsxact[17]      1.000
sigma_meds        1.000
sigma_actxmeds    1.001
shape             1.000
mu                1.000
age               1.001
age2              1.001
logDose           1.000
smplTime          1.000
medsTot           1.001
meds[1]           1.000
meds[2]           1.000
meds[3]           1.000
meds[4]           1.000
meds[5]           1.001
meds[6]           1.001
meds[7]           1.000
meds[8]           1.000
meds[9]           1.001
meds[10]          1.000
meds[11]          1.000
meds[12]          1.001
meds[13]          1.001
meds[14]          1.000
meds[15]          1.000
meds[16]          1.000
meds[17]          1.000
inhibition[1]     1.000
inhibition[2]     1.000
inhibition[3]     1.000
inhibition[4]     1.000
inhibition[5]     1.000
inhibition[6]     1.000
inhibition[7]     1.000
inhibition[8]     1.000
inhibition[9]     1.000
inhibition[10]    1.000
inhibition[11]    1.000
inhibition[12]    1.000
inhibition[13]    1.000
inhibition[14]    1.000
inhibition[15]    1.000
inhibition[16]    1.000
```

```
inhibition[17]    1.001
inhibition_PM[1]  1.000
inhibition_PM[2]  1.000
inhibition_PM[3]  1.000
inhibition_PM[4]  1.001
inhibition_PM[5]  1.000
inhibition_PM[6]  1.000
inhibition_PM[7]  1.000
inhibition_PM[8]  1.000
inhibition_PM[9]  1.000
inhibition_PM[10] 1.000
inhibition_PM[11] 1.000
inhibition_PM[12] 1.000
inhibition_PM[13] 1.000
inhibition_PM[14] 1.001
inhibition_PM[15] 1.000
inhibition_PM[16] 1.000
inhibition_PM[17] 1.000
lp__              1.000

Samples were drawn using NUTS(diag_e) at Fri Jun 19 16:39:59 2026.
For each parameter, n_eff is a crude measure of effective sample size,
and Rhat is the potential scale reduction factor on split chains (at
convergence, Rhat=1).
```

The table shows estimates and credibility intervals. The coefficients stand for:

- *mu*: estimated D/C ratio at 24h in PMs of average age, no co-medication, taking escitalopram 10 mg/day;
- *act*: the effect on the D/C ratio of CYP2C19 activity scores in individuals of average age in the without co-medication group, with plasma levels assessed at 24h, taking escitalopram 10 mg/day;
- *sex*: the effect of sex on clearance;
- *age*, *age2*: the linear and quadratic effect of age in PMs;
- *logDose*: the effect of log dose in PMs;
- *smplTime*: the effect of the log time of the sample in PMs;
- *medsTot*: the average effect on clearance of one additional co-medication in PMs;
- *meds*: the departures from the average effect given by adding an individual medication (of the 17 in the database);
- *agexact*, *age2xact*: the interaction between age (linear and quadratic) and activity scores;
- *dosexact*: the interaction between log dose and activity scores;
- *timexact*; the interaction between log sampling time and activity scores;
- *medsTotxact*: the average interaction between adding one co-medication and the activity scores, i.e. the average change in the activity score coefficient for one co-medication added;
- *medsxact*: the departure from the average interaction of adding one co-medication and the activity score for each medication in the database (17 in all);
- *inhibition_PM*: the estimated inhibition effect in PMs for each medication in the database (given by the sum of average inhibition and departures to this average due to specific inhibitions of drugs, *medsTot* + *meds*);
- *inhibition*: the estimated inhibition on CYP2C19 for each medication in the database (given by the sum of the average interaction of a co-medication with the activity scores and the individual departures from this interaction, *medsTotxact* + *medsxact*, because the *medsxact* coefficients code the departure from the mean effect of adding one co-medication).

The most interesting aspect of this models are the estimates of the individual medications. In short, “inhibition” gives the estimates and the intervals of the interaction between individual co-medications and the activity score (phenoconversion), and “inhibition_PM” the estimates of the inhibition at the PM (inhibition of other DMEs) for the inhibition of the metabolism of escitalopram.

#### Changes in metabolism due to inhibition of Escitalopram

We now turn to the estimate of the inhibition of CYP2C19.

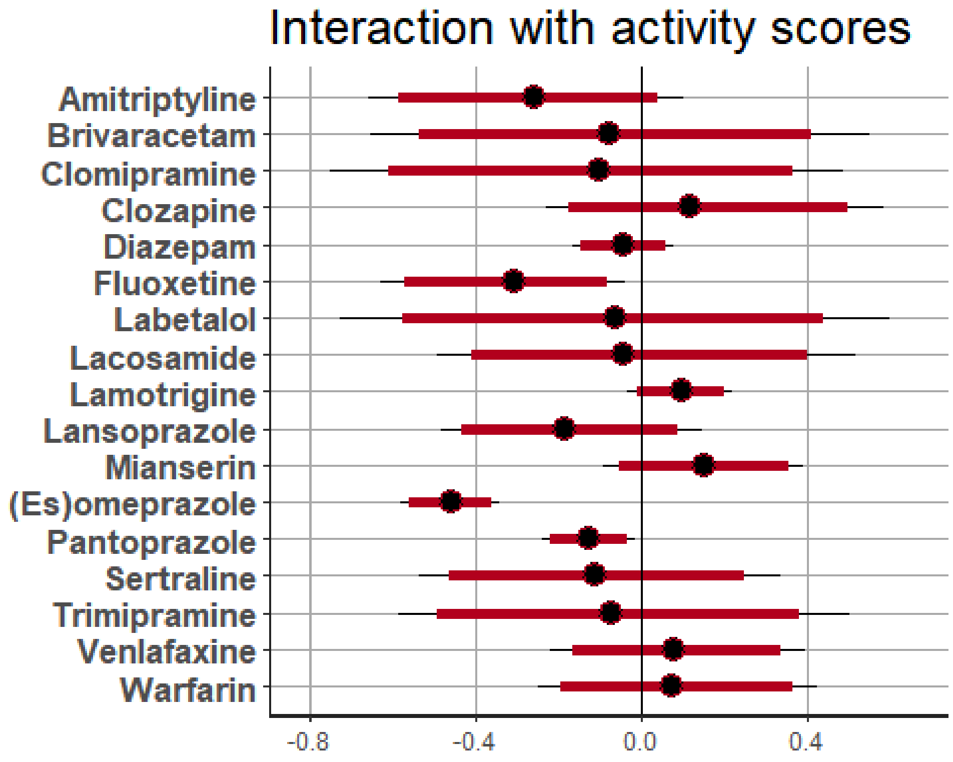


One can see clear evidence for inhibition of CYP2C19 for co-medication with fluoxetine, omeprazole, and pantoprazole (although in this latter case the inhibition is of smaller magnitude). One can see that the credibility intervals depend on the sample size for each co-medication. Starting from a dataset of about 200 observations, the amount of inhibition can be estimated up to a precision of about 10-15%.

#### Individual plots

Individual plots for selected substrates exemplify the estimated flattening of the relationship between activity scores and inhibition of metabolism of escitalopram. In case of fluoxetine, where we have few observations, the estimate shrinks the effect towards the mean effect of medications. In the plot above, we saw that fluoxetine has relatively large credibility intervals.

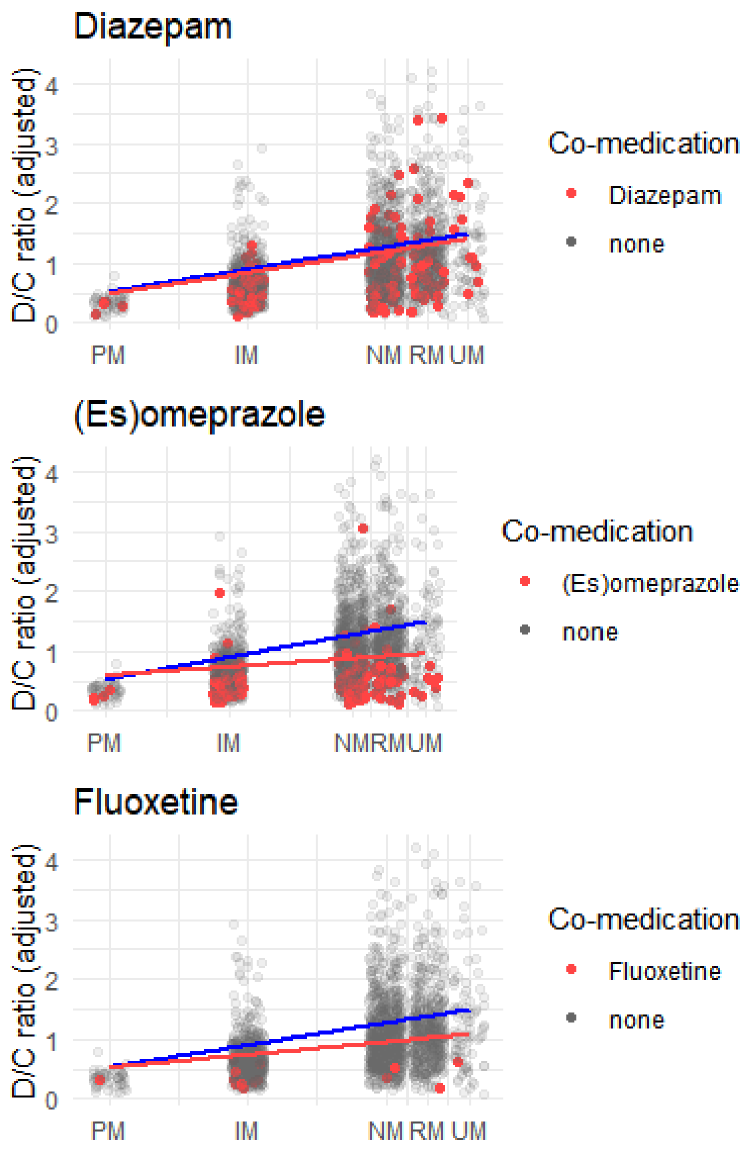


#### Correlation with pharmacogenetic data from the literature

We now investigate the association between these estimates of inhibition of CYP2C19 and *in vivo* estimates of the CYP2C19 fractional contribution to the metabolism of individual co-medication from the pharmacogenetic literature. We do this by estimating a linear model with the estimates of CYP2C19 inhibition of the previous model as outcome variables, the *in vivo* fractional contributions from the literature as predictors (Dücker & Brockmöller 2020), and the inverse of the squared standard errors of the inhibition estimates as the weights of the regression.

```
Call:
lm(formula = mean ~ FC, data = inhdata, weights = weights)

Weighted Residuals:
```

```
    Min      1Q  Median      3Q     Max 
-3.4555 -0.2813 -0.0965  0.3498  2.1759 

Coefficients:
            Estimate Std. Error t value Pr(>|t|)    
(Intercept)  0.12682    0.06354   1.996 0.064431 .  
FC          -0.46491    0.10548  -4.408 0.000509 ***
---
Signif. codes:  0 '***' 0.001 '**' 0.01 '*' 0.05 '.' 0.1 ' ' 1

Residual standard error: 1.266 on 15 degrees of freedom
Multiple R-squared:  0.5643,	Adjusted R-squared:  0.5353 
F-statistic: 19.43 on 1 and 15 DF,  p-value: 0.0005089
```

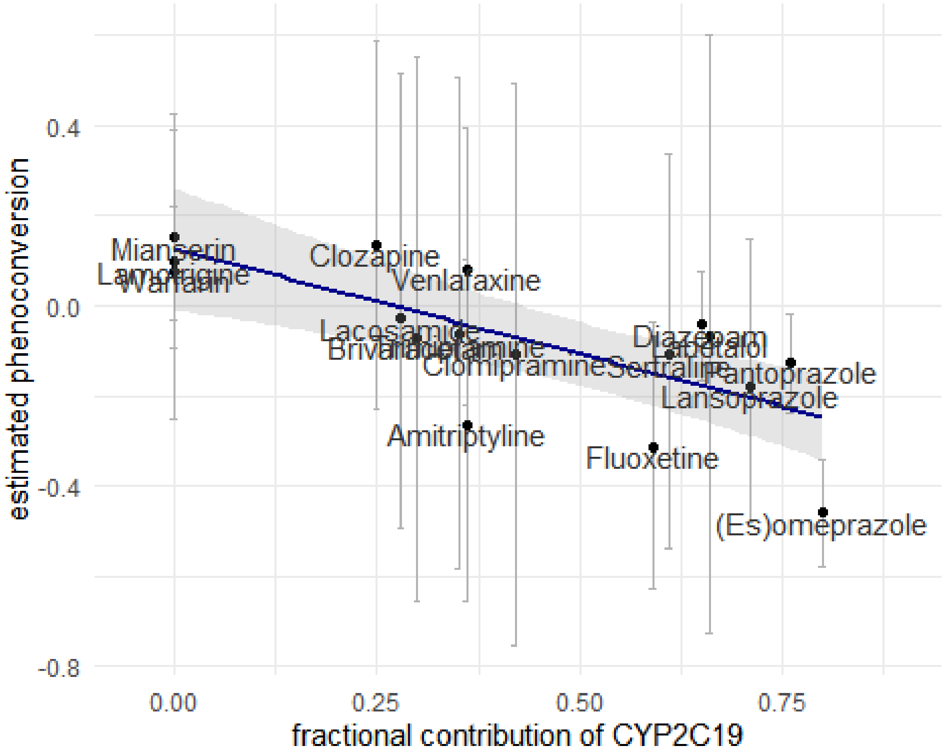


We find that the two datasets gave correlated estimates. The pharmacogenetic data explain over 55% of the variance of the competitive inhibition measured in the TDM data. Hence, estimates of inhibition of escitalopram metabolism may be used to predict the fractional contribution of CYP2C19 to the metabolism of the co-medication, and vice-versa. These figures do not account for the lower capacity of the model to predict the inhibition due to fluoxetine and omeprazole, which is not due to competitive inhibition.

In the figure, we see that co-medications with few observations cluster with large credibility intervals in the middle of the regression line. The regression is carried by the values at the extremes of the dataset. To verify the sensitivity of the analysis, we repeated it after excluding fluoxetine and omeprazole (non-competitive high inhibtion) and the co-medication with fractional clearance contribution of zero (at the other end of the inhibition specturm).

```
Call:
lm(formula = mean ~ FC, data = inhdata, subset = FC > 0 & mean >
    -0.25, weights = weights)

Weighted Residuals:
    Min      1Q  Median      3Q     Max
-0.5040 -0.3749 -0.2249  0.1747  0.5477

Coefficients:
            Estimate Std. Error t value Pr(>|t|)
(Intercept)  0.16309    0.06468   2.521  0.03269 *
FC          -0.36621    0.09735  -3.762  0.00447 **
---
Signif. codes:  0 '***' 0.001 '**' 0.01 '*' 0.05 '.' 0.1 ' ' 1

Residual standard error: 0.3965 on 9 degrees of freedom
Multiple R-squared:  0.6112,    Adjusted R-squared:  0.568
F-statistic: 14.15 on 1 and 9 DF,  p-value: 0.004474
```

While much diminished, the regression is still significant.

Here is the table of the pharmacogenetic data that went into this model.

```
        Substrate in.vivo.inhib. in.vivo.FC
1   Amitriptyline    -0.26309596       0.36
2    Brivaracetam    -0.06961630       0.30
3    Clomipramine    -0.10672773       0.42
4       Clozapine     0.13575568       0.25
5        Diazepam    -0.04117005       0.65
6      Fluoxetine    -0.31177690       0.59
7       Labetalol    -0.06621777       0.66
8      Lacosamide    -0.02792932       0.28
9     Lamotrigine     0.09707695       0.00
10   Lansoprazole    -0.17757115       0.71
11      Mianserin     0.15271602       0.00
12 (Es)omeprazole    -0.45855660       0.80
13   Pantoprazole    -0.12489225       0.76
14     Sertraline    -0.10935616       0.61
15   Trimipramine    -0.06448957       0.35
16    Venlafaxine     0.08171925       0.36
17       Warfarin     0.07869341       0.00
```

#### Inhibition of enzymes other than CYP2C19

Below, the effects in PMs are shown (recall that these reflect possible effects on DMEs other than CYP2C19 metabolizing escitalopram). No substance was detected as individually significantly affecting escitalopram clearance by other enzymes in PMs. However, pantoprazole and diazepam were possible candidates for inhibiting enzymes other than CYP2C19 compared to the other substances. Note, however, that the estimated metabolic activity in participants with co-medication was always less than in participants without. The model separates this inhibition from that attributed to CYP2C19.

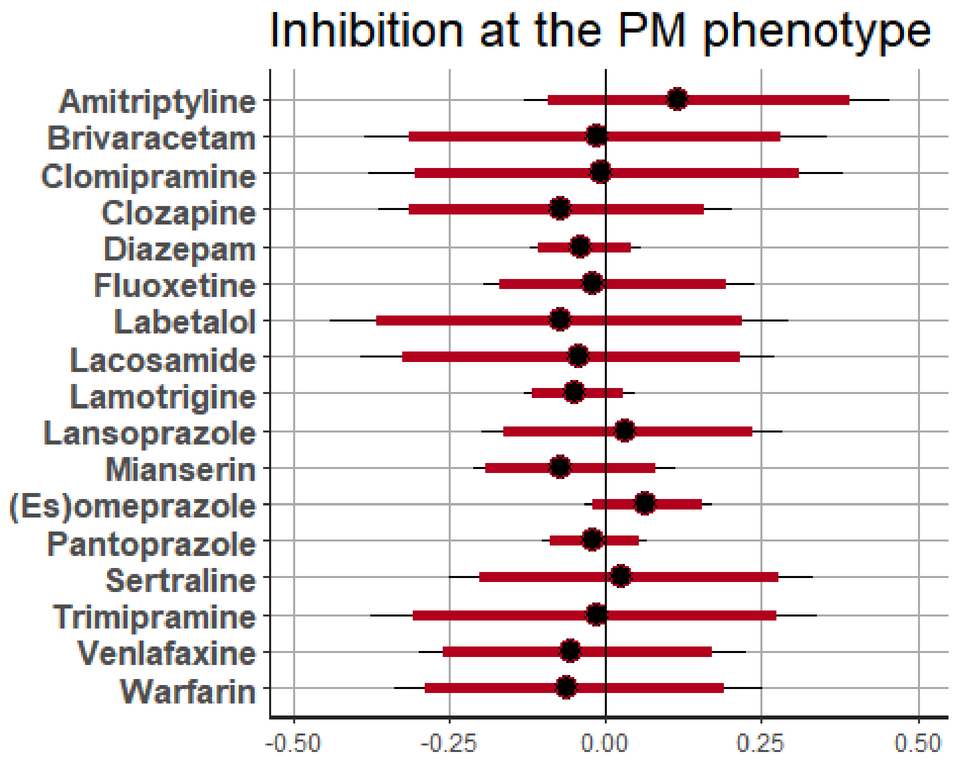
Inhibition at the PM phenotype
Amitriptyline
Brivaracetam
Clomipramine
Clozapine
Diazepam
Fluoxetine
Labetalol
Lacosamide
Lamotrigine
Lansoprazole
Mianserin
(Es)omeprazole
Pantoprazole
Sertraline
Trimipramine
Venlafaxine
Warfarin
-0.50
-0.25
0.00
0.25
0.50
